# Assessing GPT-3.5 and GPT-4 in Generating International Classification of Diseases Billing Codes

**DOI:** 10.1101/2023.07.07.23292391

**Authors:** Ali Soroush, Benjamin S. Glicksberg, Eyal Zimlichman, Yiftach Barash, Robert Freeman, Alexander W. Charney, Girish N Nadkarni, Eyal Klang

## Abstract

**Background:** Large Language Models (LLMs) like GPT-3.5 and GPT-4 are increasingly entering the healthcare domain as a proposed means to assist with administrative tasks. To ensure safe and effective use with billing coding tasks, it is crucial to assess these models’ ability to generate the correct International Classification of Diseases (ICD) codes from text descriptions.

**Objectives:** We aimed to evaluate GPT-3.5 and GPT-4’s capability to generate correct ICD billing codes, using the ICD-9-CM (2014) and ICD-10-CM and PCS (2023) systems.

**Methods:** We randomly selected 100 unique codes from each of the most recent versions of the ICD-9-CM, ICD-10-CM, and ICD-10-PCS billing code sets published by the Centers for Medicare and Medicaid Services. Using the ChatGPT interface (GPT-3.5 and GPT-4), we prompted for the ICD codes that corresponding to each provided code description. Outputs were compared with the actual billing codes across several performance measures. Errors were qualitatively and quantitatively assessed for any underlying patterns.

**Results:** GPT-4 and GPT-3.5 demonstrated varied performance across each ICD system. In ICD-9-CM, GPT-4 and GPT-3.5 achieved an exact match rate of 22% and 10%, respectively. 13% (GPT-4) and 10% (GPT-3.5) of generated ICD-10-CM codes were exact matches. Notably, both models struggled considerably with the procedurally focused ICD-10-PCS, with neither GPT-4 or GPT-3.5 producing any exactly matched codes. A substantial number of incorrect codes had semantic similarity with the actual codes for ICD-9-CM (GPT-4: 60.3%, GPT-3.5: 51.1%) and ICD-10-CM (GPT-4: 70.1%, GPT-3.5: 61.1%), in contrast to ICD-10-PCS (GPT-4: 30.0%, GPT-3.5: 16.0%).

**Conclusion:** Our evaluation of GPT-3.5 and GPT-4’s proficiency in generating ICD billing codes from ICD-9-CM, ICD-10-CM and ICD-10-PCS code descriptions reveals an inadequate level of performance. While the models appear to exhibit a general conceptual understanding of the codes and their descriptions, they have a propensity for hallucinating key details, suggesting underlying technological limitations of the base LLMs. This suggests a need for more rigorous LLM augmentation strategies and validation prior to their implementation in healthcare contexts, particularly in tasks such as ICD coding which require significant digit-level precision.

## Introduction

The International Classification of Diseases (ICD) terminology is the most widely used administrative coding system in the world (1–3). It provides a standardized representation of medical conditions and procedures, playing a critical role in clinical recordkeeping, public health surveillance, research, and billing (2).

Large Language Models (LLMs), such as GPT-3.5 and its more advanced successor GPT-4, have shown remarkable and varied capabilities that have potential to impact many domains (4–6). LLMs leverage the power of state-of-the-art deep learning algorithms, trained on extensive textual data (6). These models are capable of impressive feats, ranging from correctly answering medical board exam questions to writing poetry (4, 7). In the realm of healthcare, they could potentially support clinicians by automating administrative tasks, offering decision support, or communicating with patients (8–10).

These models have an ability to create text that appears human-generated, offering hope that this technological advancement will unlock important downstream natural language processing tasks like assigning ICD codes based on unstructured medical text descriptions. However, the accuracy of LLMs for administrative tasks in medicine has not been thoroughly assessed yet. A prominent concern is the models’ propensity to ’hallucinate,’ i.e., generate plausible sounding, but factually incorrect information (11–14). Before LLMs can be used to automate burdensome administrative tasks like assigning ICD billing codes based on clinical documentation, the propensity for hallucination must be delineated and, ultimately, mitigated.

Our study analyzes how GPT-3.5 and GPT-4 fare in the context of ICD billing code generation. We systematically assess their performance in matching ICD billing codes to their descriptions across multiple versions of the classification system.

## Methods

### ICD Code Selection

We obtained most recent lists of ICD-9-CM-CM (2014), ICD-10-CM (2023), and ICD-10-PCS (2023) billing codes from the Centers for Medicare & Medicaid Services (CMS)(15). From each list, we randomly selected 100 unique codes, leading to a total of 300 ICD billing codes for our dataset.

### LLMs Used

For this study, we deployed two commercially available Large Language Models (LLMs): GPT-3.5 and GPT-4, both of which were developed by OpenAI (5, 16). The underlying data used to train these models has not been publicly released, but presumably contains a combination of all publicly available online data as well as private datasets. Their data is inclusive to September 2021.

### ICD Code Generation

We used the public ChatGPT interface to produce the corresponding ICD billing code for each of the 300 code descriptions. We used the following natural language prompt, where <ICD coding system> refers to ICD-9-CM-CM, ICD-10-CM, or ICD-10-PCS: *“Dear GPT, we will feed you a list of <ICD coding system> descriptions. Please write the matching <ICD coding system> codes. Please, return in a table format.”* The descriptions were provided to the LLMs in batches of 10 per prompt to improve processing efficiency.

### Performance Evaluation

To assess LLM billing code generation performance, we determined the number of exact code matches, billable codes, nonbillable codes, and nonexistent codes for the output codes. Nonbillable codes are generally more non-specific than billing codes. We labeled exact matches by comparing each output code with the original code for its originating description. We labeled billable codes and obtained their descriptions by matching output codes with any code from the same coding system present in the original CMS billing code lists. To label the nonbillable codes and obtain their descriptions, we applied the UMLS Metathesaurus (17) to the remaining unmatched codes. Any remaining codes without an assigned description were considered to be nonexistent codes. For all generated codes, we assessed *semantic* and *syntactic* similarity measures to interrogate the LLMs’ broader understanding of the ICD coding systems. To assess semantic similarity, two physicians (EK, AS) assessed for any meaningful conceptual similarity between a generated billable code’s description and its original code description. In terms of syntactic similarity, we ascertained if a generated code differed from its original code by a single character or less, including differences in code length. Syntactic and semantic similarity percentages were calculated by dividing each similarity count by the total number of generated codes.

### Error Analysis

We conducted error analyses for each coding system to identify with which specific contexts the models exceled or struggled. We first measured the semantic and syntactic similarities of valid billing codes that did not match with their original codes. To assess the effect of code and description complexity on model performance, we assessed the relationships between performance and code length and description length respectively. We could not assess this for ICD-10-PCS due to low number of correctly matched codes (3 for GPT-4 and 0 for GPT-3.5). We additionally performed a qualitative analysis of the nature and context of errors, focusing on how well the models handled unspecified conditions, complex conditions, and semantic variations in descriptions. Two physicians performed this analysis using a consensus approach.

### Statistical Analysis

We summarized the ICD code generating performance for each model using descriptive statistics. We calculated counts and percentages for exact code matches, billable codes, and nonbillable codes exact match counts and percentages for each score across the ICD versions. Counts and percentages were similarly calculated for semantic and syntactic similarity. We used Fisher’s exact test to compare each performance metric between GPT-4 with GPT-3.5. To assess the relationships between exact matches and code and description length respectively, we applied the Mann-Whitney U test. We reported the median and interquartile range (IQR) values for the lengths for each case. We considered a p-value less than 0.05 statistically significant, indicating a meaningful performance difference between the two models on a given ICD code dataset. We coded all analyses in Python (Version 3.9.16).

## Results

### Performance Evaluation

When assessing the ICD code generation performance of GPT-3.5 and GPT-4 from our predefined list of code descriptions (**Supplementary tables 1-3**), we found variable results across the different ICD coding systems (**Table 1**). For ICD-9-CM, GPT-4 generated exactly matched codes for 22% of cases, billable codes for 72% of cases, nonbillable codes for 26% of cases, and nonexistent codes for 2% of cases. GPT-3.5 generated a significantly lower rate of exactly matched codes (10%, p=0.033), but had similar rates of billable (76%, p=0.629), nonbillable (20%, p=0.401), and nonexistent codes (4%, p=0.683). For the ICD-10-CM system, both models produced fewer exact match and nonbillable codes and more nonexistent codes. GPT-4 generated exactly matched codes for 13% of cases, billable codes for 77% of cases, nonbillable codes for 3% of cases, and nonexistent codes for 20% of cases. In comparison to GPT-4, GPT-3.5 generated similar rates of exactly matched (5%, p=0.081), billable (67%, p=0.156), nonbillable (4%, p=1.000), and nonexistent codes (28%, p=0.246). GPT-4 and GPT-3.5 both had the lower ICD code generation performance for the ICD-10-PCS system, with both producing no exact match codes or nonbillable codes. GPT-4 was able to generate billable codes for 39% of cases, with the remainder (61%) of the codes being nonexistent GPT-3.5 had similar rates for billable (30%, p=0.234) and nonexistent (70%, p=0.234) codes.

**Table 1:**
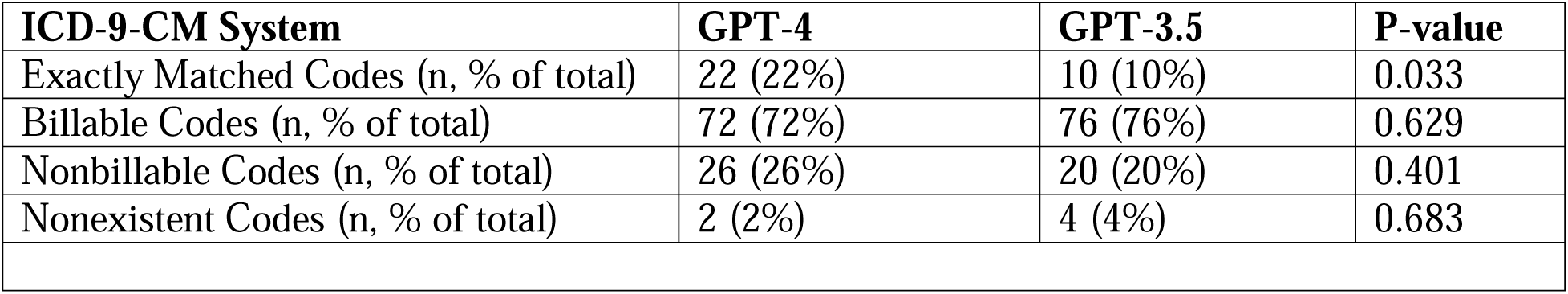
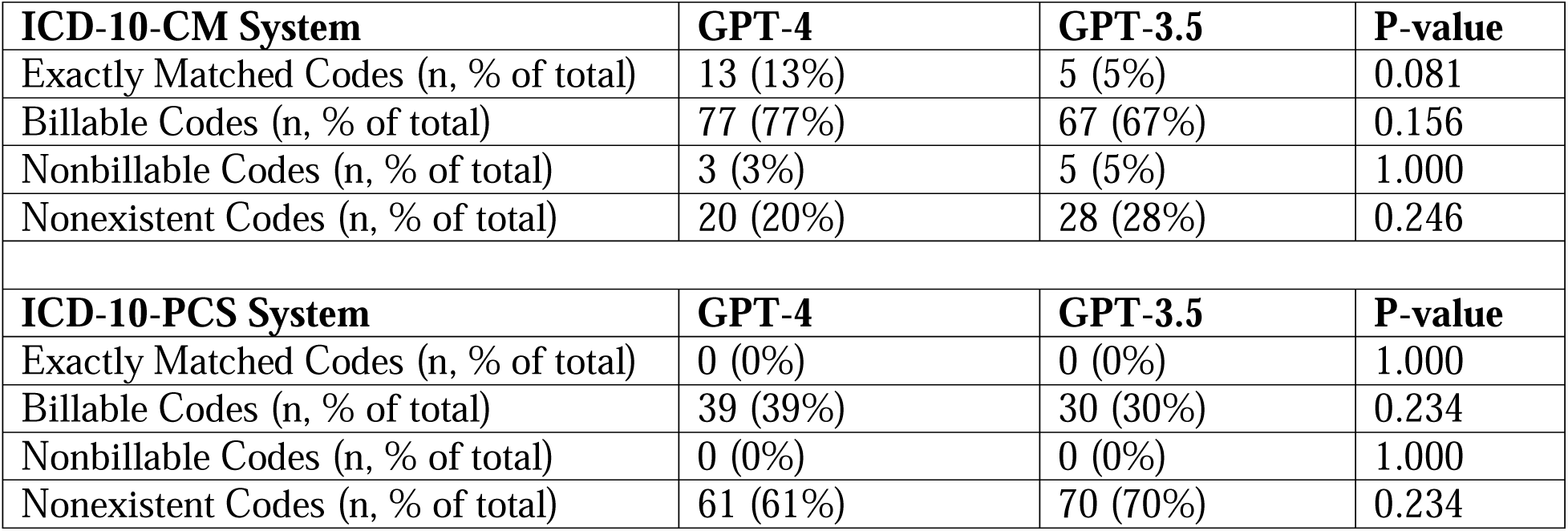
Validity and Accuracy of GPT-generated ICD codes; abbreviations: ICD International Classification of Diseases

### Semantic and syntactic similarity

**Table 2** reveals that both models demonstrated a high level of semantic, or conceptual, similarity for all three ICD systems, though GPT-4 outperformed GPT-3.5. When assessing ICD-9-CM, GPT-4 achieved a semantic similarity of 60%) versus 43% for GPT 3.5 (p=0.003). In the case of ICD-10-CM, GPT-4 and GPT-3.5 both attained higher semantic similarities (74% and 63% respectively) than they did for ICD-9-CM. However, both models struggled with the ICD-10-PCS system, showing semantic similarity for only 30% (GPT-4) and 16% (GPT-3.5) of the codes.

**Table 2:** Semantic and syntactic similarity of billable GPT-generated ICD codes for all generated codes; abbreviations: ICD International Classification of Diseases

| **ICD-9-CM** | **GPT-4** | **GPT-3.5** | **P-value** |
| --- | --- | --- | --- |
| Syntactically Similar Codes (n, %) | 60 (60%) | 43 (43%) | 0.003 |
| Semantically Similar Codes (n, %) | 69 (69%) | 56 (56%) | 0.002 |
| **ICD-10-CM** | **GPT-4** | **GPT-3.5** | **P-value** |
| Syntactically Similar Codes (n, %) | 36 (36%) | 19 (19%) | 0.007 |
| Semantically Similar Codes (n, %) | 74 (74%) | 63 (63%) | 0.008 |
| **ICD-10-PCS** | **GPT-4** | **GPT-3.5** | **P-value** |
| Syntactically Similar Codes (n, %) | 3 (3%) | 0 (0.0%) | 0.240 |
| Semantically Similar Codes (n, %) | 30 (30%) | 16 (16%) | 0.003 |

Rates of syntactic, or character-level, code similarity were more variable. GPT-4 displayed a high degree of syntactic similarity when generating ICD-9-CM codes (60%), but this measure dropped greatly for ICD-10-CM (36%) and even more so for ICD-10-PCS (3%). GPT-3.5 demonstrated a similar, but consistently lower scoring pattern of syntactic similarity scores across all three ICD terminologies, with rates of 43% for ICD-9-CM, 19% for ICD-10-CM, and 0% for ICD-10-PCS.

### Error analysis

When repeating the similarity analysis on only the incorrect codes, we found that similar rates of semantic similarity as in the overall analysis (**Table 3**). For the codes produced by GPT-4, 60.3% of incorrect ICD-9-CM codes, 70.1% of incorrect ICD-10-CM codes, and 30% of incorrect ICD-10-PCS codes were semantically similar. GPT-3.5 produced a similar semantic similarity pattern, but with consistently lower scores: 51.1% for ICD-9-CM, 61.1% for ICD-10-CM, and 16.0% for ICD-10-PCS.

**Table 3:** Semantic and syntactic similarity of GPT-generated billable GPT-generated ICD codes for all incorrectly generated codes; abbreviations: ICD International Classification of Diseases

| **ICD-9-CM** | **GPT-4** | **GPT-3.5** | **P-value** |
| --- | --- | --- | --- |
| Syntactically Similar Codes (n, %) | 38 (48.7%) | 33 (36.7%) | 0.121 |
| Semantically Similar Codes (n, %) | 47 (60.3%) | 46 (51.1%) | 0.277 |
| **ICD-10-CM** | **GPT-4** | **GPT-3.5** | **P-value** |
| Syntactically Similar Codes (n, %) | 23 (26.4%) | 14 (14.7%) | 0.065 |
| Semantically Similar Codes (n, %) | 60 (70.1%) | 58 (61.1%) | 0.215 |
| **ICD-10-PCS** | **GPT-4** | **GPT-3.5** | **P-value** |
| Syntactically Similar Codes (n, %) | 3 (3.0%) | 0 (0.0%) | 0.246 |
| Semantically Similar Codes (n, %) | 30 (30.0%) | 16 (16.0%) | 0.028 |

Our error analysis unveiled several findings. With all code systems, the models tended toward generating more general nonbillable codes for complex and lengthy code descriptions such as case 1 in **Supplementary Table 1** (“Other open skull fracture with cerebral laceration and contusion, with moderate [1-24 hours] loss of consciousness”, ICD-9-CM code 80363) other similar cases 11, 14, 20. **Table 4** presents quantitative analyses of the associations between model exact match performance and code length and description length respectively. Overall, code and description length were inversely associated with exact matches. Significant relationships were present for ICD-9-CM descriptions lengths and GPT-4 and GPT-3.5 exact match performance, ICD-10-CM code lengths and GPT-4 and GPT-3.5 exact match performance, and ICD-10-CM description length and GPT-4 exact match performance.

**Table 4:** ICD-9-CM and ICD-10-CM length of codes and descriptions association with exact matches (Mann Whitney U test); abbreviations: ICD International Classification of Diseases, IQR interquartile range

| **ICD-9-CM** | **GPT-4** |  |  | **GPT-3.5** |  |  |
| --- | --- | --- | --- | --- | --- | --- |
|  | **Exact Matches** | **Not Exact Matches** | **P-value** | **Exact Matches** | **Not Exact Matches** | **P-value** |
| Code length (median, IQR) | 4.0 (4.0 - 5.0) | 4.0 (4.0 - 5.0) | 0.917 | 5.0 (4.0 - 5.0) | 4.0 (4.0 - 5.0) | 0.410 |
| Description length (median, IQR) | 38.0 (26.5 - 54.0) | 46.0 (35.0 - 75.0) | 0.081 | 27.5 (23.5 - 52.5) | 46.5 (35.0 - 74.5) | 0.025 |
| **ICD-10-CM** | **GPT-4** |  |  | **GPT-3.5** |  |  |

|  | **Exact Matches** | **Not Exact Matches** | **P-value** | **Exact Matches** | **Not Exact Matches** | **P-value** |
| --- | --- | --- | --- | --- | --- | --- |
| Code length (median, IQR) | 5.0<br>(4.0 - 7.0) | 7.0<br>(7.0 - 7.0) | < 0.001 | 5.0<br>(4.0 - 5.0) | 7.0<br>(6.0 - 7.0) | 0.001 |
| Description length (median, IQR) | 49.0<br>(38.0 - 63.0) | 84.0<br>(62.0 - 112.5) | <0.001 | 63.0<br>(38.0 - 77.0) | 80.0<br>(59.5 - 111.0) | 0.131 |

Certain condition categories consistently had worse exact match accuracy and semantic similarity scores, driven again by a tendency for the LLMs to rely on non-billing codes for complex scenarios. For example, generic non-billable codes were generated for pregnancy related disorders such as cases 9 (“Major puerperal infection”, ICD-9-CM code 67082) and 20 (“Mild or unspecified pre-eclampsia, unspecified as to episode of care”, ICD-9-CM code 64224). Even when the models generated billable codes, the codes often differed in severity or specificity, as observed in ICD-9-CM case 5 (“Injury to hypoglossal nerve”, ICD-9-CM code 9517) and others (7, 12, 32, 33, 50, 62).

In the context of ICD-10-CM code generation (**Supplemental Table 2**), GPT-4 had trouble achieving the high level of specificity required by this coding system, as only 14 generated codes were exact matches. Nevertheless, the overall level of specificity was generally higher than with ICD-9-CM, as demonstrated by case 7 (“Acute embolism and thrombosis of unspecified deep veins of right lower extremity”, ICD-10-CM code I82401), case 15 (“Benign carcinoid tumor of the transverse colon”, ICD-10-CM code D3A023), and others (14, 27, 34, 46). The low number of non-billable codes (3%) generated and high rate of semantic similarity for generated codes supports the overall trend toward specificity for generated ICD-10-CM codse. Incorrectly assigned billable codes had errors related to highly specific features of the description such as injury type, laterality, complexity, and complications (e.g. cases 1, 14, 41, 75). Nearly all valid codes were aligned with the temporal features of the code (initial vs subsequent encounter), which is represented by the last letter in the ICD-10-CM code. Nonexistent codes were most frequent for codes related to maternal care (0/2, 0.0%), vehicle-related injuries (4/8, 50.0%), and joint-related conditions (4/7, 57.1%).

For ICD-10-PCS, GPT-4 was unable to correctly match any codes. However, the model was able to achieve some semantic similarity for 30% of generated codes. These codes differed in much more in terms of anatomical locations, laterality, maneuvers, and procedures. There was a high degree of concordance for procedure approach, which is represented by the last letter in the ICD-10-PCS code.

## Discussion

Our study is the first to evaluate the ICD billing code mapping performance of both GPT-3.5 and GPT-4. Their underlying LLM technology holds the promise of automating the mapping of these core administrative terminologies in healthcare, with significant implications for billing, clinical decision making, quality improvement, research, and health policy.

The findings revealed that GPT-4 generally outperformed GPT-3.5 in generating exact match ICD billing codes, billing codes, and non-billing codes though overall performance was not reliable across both. Performance varied considerably across different versions of the ICD system. Both models showed some success with ICD-9-CM, but struggled more with the newer ICD-10-CM and ICD-10-PCS systems. This reflects the increased complexity, granularity, and comprehensiveness of these more recent versions (18) and highlights the difficulty faced by LLMs in handling a critical healthcare-related task.

Interestingly, despite their struggle with exact code generation, the models often generated codes that were semantically or syntactically similar to the actual codes. Both models demonstrated a better grasp of the more commonly used and queried ICD-9-CM and ICD-10-CM diagnosis code systems compared to the ICD-10-PCS procedure code system. This suggests that the base versions of these LLMs can parse the general descriptive nature of ICD codes, but could not attain precision, potentially due to a tendency to generalize or, worse, hallucinate data when the input is sparse or ambiguous (6, 12). The performance differences across systems may be due to the broader availability of ICD-9-CM and ICD-10-CM coding references in the LLM training data (19), which is drawn from publicly available web and print data. Alternatively, this behavior may be due to the underlying architecture of the LLMs we tested, as they have been trained to produce generalized responses for a public audience, rather than exact and technical responses, unless specified otherwise (5, 16, 20).

Our formal error analysis highlighted additional patterns of poor ICD billing code generation performance. Complex and lengthy descriptions, as well as certain condition categories, proved challenging for the model to parse. It should be noted that nonbillable codes, shorter codes, and shorter descriptions usually reflect more generalized conditions. These patterns of poor performance must be addressed before GPT-based LLMs can be used to interface with ICD terminologies for billing purposes. Additional prompt engineering to encourage exact matching of ICD billing codes may be able to improve accuracy (21).

The inability of LLMs to parse ICD codes consistently has parallels with other previously identified general technical limitations of these models. Due to the nature of how LLMs algorithmically break up text into “tokens”, some have difficulty with tasks that require an understanding the structure of information contained within each token (22–24). Similarly, LLMs are unable to consistently reverse the spelling of words, perform math problems, or complete complex coding tasks without the use of external software or model fine-tuning. When we evaluated ICD codes in OpenAI’s tokenization tool, we observed that tokenization behavior was not consistent with the underlying ICD terminology structures, leaving the models without key digit-level hierarchical information during the model training process (25, 26). To address this technical issue, an GPT-3.5 or GPT-4 ICD tool will need to be linked to additional software layers that can facilitate an interface between LLMs and ICD billing codes (11, 27–32).

This study’s results reaffirm the tendency for LLMs to produce realistic-appearing, but incorrect information—a significant obstacle to their use in the healthcare setting. While this study showed that such “hallucination” behavior did not significantly impact broad semantic understanding, it often compromised precision to a degree that is not acceptable for medical coding purposes (33).

As LLM technology is increasingly embedded into healthcare, understanding these limitations is crucial. LLM accuracy while interfacing with ICD terminologies must be improved before they can help realize their potential in streamlining administrative tasks, supporting clinicians, and improving patient care. Additional LLM performance enhancement strategies such as prompt engineering, database linkage, model fine-tuning, and others can potentially address this in the near future.

### Limitations

Our study has a few limitations. First, we used a sample of conditions and procedures for testing, which may not represent the codes most frequently used in real-world coding scenarios. Second, we did not evaluate advanced LLM performance enhancement strategies such as prompt engineering, database-linkage, or model fine-tuning. Lastly, we did not evaluate the models’ performance in the context of real-world clinical narratives, which often involves complex and ambiguous language that does not always have a clear ground-truth mapping.

### Conclusion

Our evaluation of baseline GPT-3.5 and GPT-4’s proficiency in generating ICD billing codes from the ICD-9-CM, ICD-10-CM and ICD-10-PCS coding systems reveals an inadequate level of performance for downstream use cases. While the models do exhibit an understanding of the conditions, as demonstrated by the generation of codes semantically related to the actual ones, they tend to exhibit a propensity for data hallucination, producing codes that are not entirely accurate. This suggests a need for rigorous validation and refinement processes prior to their implementation in healthcare contexts, particularly in tasks such as ICD billing coding which require precision. Further, the integration with external tools may be necessary to enhance their performance.

## Data Availability

All data produced in the present study are available upon reasonable request to the authors.

## Data Availability Statement

The data that support the findings of this study are available from the corresponding author, AS, upon reasonable request.

## Ethics Approval

Since all data and responses were publicly available, approval from the institutional review board was not sought.

## References

1. Organization WH. History of the development of the ICD [June 19, 2023]. Available from: https://cdn.who.int/media/docs/default-source/classification/icd/historyoficd.pdf.

2. Organization WH. Importance of ICD. Available from: https://www.who.int/standards/classifications/frequently-asked-questions/importance-of-icd.

3. Wood PH. Applications of the International Classification of Diseases. World Health Stat Q. 1990;43(4):263–8. PubMed PMID: 2293495.

4. Bubeck S, Chandrasekaran V, Eldan R, Gehrke J, Horvitz E, Kamar E, et al. Sparks of Artificial General Intelligence: Early experiments with GPT-42023 March 01, 2023:[arXiv:2303.12712 p.]. Available from: https://ui.adsabs.harvard.edu/abs/2023arXiv230312712B.

5. OpenAI. GPT-4 Technical Report2023 March 01, 2023:[arXiv:2303.08774 p.]. Available from: https://ui.adsabs.harvard.edu/abs/2023arXiv230308774O.

6. Zhao WX, Zhou K, Li J, Tang T, Wang X, Hou Y, et al. A Survey of Large Language Models2023 March 01, 2023:[arXiv:2303.18223 p.]. Available from: https://ui.adsabs.harvard.edu/abs/2023arXiv230318223Z.

7. Nori H, King N, McKinney SM, Carignan D, Horvitz E. Capabilities of GPT-4 on Medical Challenge Problems2023 March 01, 2023:[arXiv:2303.13375 p.]. Available from: https://ui.adsabs.harvard.edu/abs/2023arXiv230313375N.

8. Lee P, Bubeck S, Petro J. Benefits, Limits, and Risks of GPT-4 as an AI Chatbot for Medicine. N Engl J Med. 2023;388(13):1233–9. doi: 10.1056/NEJMsr2214184. PubMed PMID: 36988602.

9. Patel SB, Lam K. ChatGPT: the future of discharge summaries? Lancet Digit Health. 2023;5(3):e107–e8. Epub 20230206. doi: 10.1016/S2589-7500(23)00021-3. PubMed PMID: 36754724.

10. Moor M, Banerjee O, Abad ZSH, Krumholz HM, Leskovec J, Topol EJ, et al. Foundation models for generalist medical artificial intelligence. Nature. 2023;616(7956):259–65. Epub 20230412. doi: 10.1038/s41586-023-05881-4. PubMed PMID: 37045921.

11. Manakul P, Liusie A, Gales MJF. SelfCheckGPT: Zero-Resource Black-Box Hallucination Detection for Generative Large Language Models2023 March 01, 2023:[arXiv:2303.08896 p.]. Available from: https://ui.adsabs.harvard.edu/abs/2023arXiv230308896M.

12. McKenna N, Li T, Cheng L, Hosseini MJ, Johnson M, Steedman M. Sources of Hallucination by Large Language Models on Inference Tasks2023 May 01, 2023:[arXiv:2305.14552 p.]. Available from: https://ui.adsabs.harvard.edu/abs/2023arXiv230514552M.

13. Li J, Cheng X, Zhao WX, Nie J-Y, Wen J-R. HaluEval: A Large-Scale Hallucination Evaluation Benchmark for Large Language Models2023 May 01, 2023:[arXiv:2305.11747 p.]. Available from: https://ui.adsabs.harvard.edu/abs/2023arXiv230511747L.

14. Ji Z, Lee N, Frieske R, Yu T, Su D, Xu Y, et al. Survey of Hallucination in Natural Language Generation2022 February 01, 2022:[arXiv:2202.03629 p.]. Available from: https://ui.adsabs.harvard.edu/abs/2022arXiv220203629J.

15. Services CfMM. ICD Code Lists 2022 [updated September 28, 2022]. Available from: https://www.cms.gov/medicare/coordination-benefits-recovery-overview/icd-code-lists.

16. Ouyang L, Wu J, Jiang X, Almeida D, Wainwright CL, Mishkin P, et al. Training language models to follow instructions with human feedback2022 March 01, 2022:[arXiv:2203.02155 p.]. Available from: https://ui.adsabs.harvard.edu/abs/2022arXiv220302155O.

17. Bodenreider O. The Unified Medical Language System (UMLS): integrating biomedical terminology. Nucleic Acids Res. 2004;32(Database issue):D267–70. doi: 10.1093/nar/gkh061. PubMed PMID: 14681409; PubMed Central PMCID: PMC308795.

18. Topaz M, Shafran-Topaz L, Bowles KH. ICD-9 to ICD-10: evolution, revolution, and current debates in the United States. Perspect Health Inf Manag. 2013;10(Spring):1d. Epub 20130401. PubMed PMID: 23805064; PubMed Central PMCID: PMC3692324.

19. Razeghi Y, Logan RL, IV, Gardner M, Singh S. Impact of Pretraining Term Frequencies on Few-Shot Reasoning2022 February 01, 2022:[arXiv:2202.07206 p.]. Available from: https://ui.adsabs.harvard.edu/abs/2022arXiv220207206R.

20. Kenton Z, Everitt T, Weidinger L, Gabriel I, Mikulik V, Irving G. Alignment of Language Agents2021 March 01, 2021:[arXiv:2103.14659 p.]. Available from: https://ui.adsabs.harvard.edu/abs/2021arXiv210314659K.

21. White J, Fu Q, Hays S, Sandborn M, Olea C, Gilbert H, et al. A Prompt Pattern Catalog to Enhance Prompt Engineering with ChatGPT2023 February 01, 2023:[arXiv:2302.11382 p.]. Available from: https://ui.adsabs.harvard.edu/abs/2023arXiv230211382W.

22. Yuan Z, Yuan H, Tan C, Wang W, Huang S. How well do Large Language Models perform in Arithmetic tasks?2023 March 01, 2023:[arXiv:2304.02015 p.]. Available from: https://ui.adsabs.harvard.edu/abs/2023arXiv230402015Y.

23. Kim J, Hong G, Kim K-m, Kang J, Myaeng S-H, editors. Have You Seen That Number? Investigating Extrapolation in Question Answering Models2021 November; Online and Punta Cana, Dominican Republic: Association for Computational Linguistics.

24. Nogueira R, Jiang Z, Lin J. Investigating the Limitations of Transformers with Simple Arithmetic Tasks2021 February 01, 2021:[arXiv:2102.13019 p.]. Available from: https://ui.adsabs.harvard.edu/abs/2021arXiv210213019N.

25. OpenAI. Tokenizer. Available from: https://platform.openai.com/tokenizer.

26. Brown TB, Mann B, Ryder N, Subbiah M, Kaplan J, Dhariwal P, et al. Language Models are Few-Shot Learners2020 May 01, 2020:[arXiv:2005.14165 p.]. Available from: https://ui.adsabs.harvard.edu/abs/2020arXiv200514165B.

27. Peng B, Galley M, He P, Cheng H, Xie Y, Hu Y, et al. Check Your Facts and Try Again: Improving Large Language Models with External Knowledge and Automated Feedback2023 February 01, 2023:[arXiv:2302.12813 p.]. Available from: https://ui.adsabs.harvard.edu/abs/2023arXiv230212813P.

28. Huang J, Gu SS, Hou L, Wu Y, Wang X, Yu H, et al. Large Language Models Can Self Improve2022 October 01, 2022:[arXiv:2210.11610 p.]. Available from: https://ui.adsabs.harvard.edu/abs/2022arXiv221011610H.

29. Azaria A, Mitchell T. The Internal State of an LLM Knows When its Lying2023 April 01, 2023:[arXiv:2304.13734 p.]. Available from: https://ui.adsabs.harvard.edu/abs/2023arXiv230413734A.

30. Lin S, Hilton J, Evans O. Teaching Models to Express Their Uncertainty in Words2022 May 01, 2022:[arXiv:2205.14334 p.]. Available from: https://ui.adsabs.harvard.edu/abs/2022arXiv220514334L.

31. Lu P, Peng B, Cheng H, Galley M, Chang K-W, Nian Wu Y, et al. Chameleon: Plug-and-Play Compositional Reasoning with Large Language Models2023 April 01, 2023:[arXiv:2304.09842 p.]. Available from: https://ui.adsabs.harvard.edu/abs/2023arXiv230409842L.

32. Dai H, Liu Z, Liao W, Huang X, Cao Y, Wu Z, et al. AugGPT: Leveraging ChatGPT for Text Data Augmentation2023 February 01, 2023:[arXiv:2302.13007 p.]. Available from: https://ui.adsabs.harvard.edu/abs/2023arXiv230213007D.

33. The Lancet Digital H. ChatGPT: friend or foe? Lancet Digit Health. 2023;5(3):e102. Epub 20230206. doi: 10.1016/S2589-7500(23)00023-7. PubMed PMID: 36754723.

