## Supplementary Tables for "Assessing GPT-3.5 and GPT-4 in Generating International Classification of Diseases Billing Codes"

Supplementary Table 1: Code-level generated outputs and similarity for ICD-9-CM

| Case | Original ICD-9-CM code | Original ICD-9-CM code description | GPT-4 Code | Description matching GPT-4 code | Description matching billable GPT-4 code | GPT-4 code semantic similarity | GPT-4 code syntactic similarity | GPT-3.5 Code | Description matching GPT-3.5 code | Description matching billable GPT-3.5 code | GPT-3.5 code semantic similarity | GPT-3.5 code syntactic similarity |
|---|---|---|---|---|---|---|---|---|---|---|---|---|
| 1 | 80363 | Other open skull fracture with cerebral laceration and contusion, with moderate [1-24 hours] loss of consciousness | 8518 | Other and unspecified cerebral laceration and contusion, without mention of open intracranial wound | | nan | False | 80309 | Other closed skull fracture without mention of intracranial injury, with concussion, unspecified | Other closed skull fracture without mention of intracranial injury, with concussion, unspecified | Skull fracture | False |
| 2 | 6978 | Other lichen, not elsewhere classified | 6978 | Other lichen, not elsewhere classified | Other lichen, not elsewhere classified | Exact Match | True | 6979 | Lichen, unspecified | Lichen, unspecified | Lichen, unspecified | True |
| 3 | 20242 | Leukemic reticuloendotheliosis, intrathoracic lymph nodes | 2024 | Leukemic reticuloendotheliosis | | nan | True | 20288 | Other malignant lymphomas, lymph nodes of multiple sites | Other malignant lymphomas, lymph nodes of multiple sites | nan | False |
| 4 | 485 | Bronchopneumonia, organism unspecified | 485 | Bronchopneumonia, organism unspecified | Bronchopneumonia, organism unspecified | Exact Match | True | 486 | Pneumonia, organism unspecified | Pneumonia, organism unspecified | Pneumonia | True |
| 5 | 9517 | Injury to hypoglossal nerve | 9516 | Injury to accessory nerve | Injury to accessory nerve | Nerve injury | True | 9555 | Injury to cutaneous sensory nerve, upper limb | Injury to cutaneous sensory nerve, upper limb | Nerve injury | False |
| 6 | 78830 | Urinary incontinence, unspecified | 7883 | Urinary incontinence | | nan | True | 7883 | Urinary incontinence | | nan | True |
| 7 | 71842 | Contracture of joint, upper arm | 71881 | Other joint derangement, not elsewhere classified, shoulder region | Other joint derangement, not elsewhere classified, shoulder region | Upper arm injury | False | 71843 | Contracture of joint, forearm | Contracture of joint, forearm | nan | True |
| 8 | E9808 | Poisoning by arsenic and its compounds, undetermined whether accidentally or purposely inflicted | 9851 | Gonococcal synovitis and tenosynovitis; Toxic effect of arsenic and its compounds | Toxic effect of arsenic and its compounds | Arsenic toxicosis | False | 9859 | Other gonococcal infection of joint; Toxic effect of unspecified metal | Toxic effect of unspecified metal | Metal toxicosis | False |
| 9 | 67082 | Other major puerperal infection, delivered, with mention of postpartum complication | 6708 | Other major puerperal infection | | nan | True | 67029 | | | nan | False |
| 10 | 65190 | Unspecified multiple gestation, unspecified as to episode of care or not applicable | 651 | Omsk hemorrhagic fever; Multiple gestation | | nan | True | 6519 | Unspecified multiple gestation | | nan | True |
| 11 | 85133 | Cortex (cerebral) laceration with open intracranial wound, with moderate [1-24 hours] loss of consciousness | 8512 | Cortex (cerebral) laceration without mention of open intracranial wound | | nan | True | 8512 | Cortex (cerebral) laceration without mention of open intracranial wound | | nan | True |
| 12 | 8460 | Sprain of lumbosacral (joint) (ligament) | 8472 | Sprain of lumbar | Sprain of lumbar | Lumbar region | False | 846 | Intestinal infection due to other anaerobes; Malaria, unspecified; Sprains and strains of sacroiliac region | | nan | True |
| 13 | 44321 | Dissection of carotid artery | 44321 | Dissection of carotid artery | Dissection of carotid artery | Exact Match | True | 44329 | Dissection of other artery | Dissection of other artery | Artery dissection | True |
| 14 | 66992 | Unspecified complication of labor and delivery, delivered, with mention of postpartum complication | 6699 | Unspecified complication of labor and delivery | | nan | True | 6699 | Unspecified complication of labor and delivery | | nan | True |
| 15 | 1280 | Other specified respiratory tuberculosis, unspecified | 119 | Unspecified pulmonary tuberculosis | | nan | False | 118 | Other specified pulmonary tuberculosis; Opportunistic mycoses | Opportunistic mycoses | nan | True |
| 16 | V301 | Single liveborn, born before admission to hospital | V3000 | Single liveborn, born in hospital, delivered without mention of cesarean section | Single liveborn, born in hospital, delivered without mention of cesarean section | Birth event | True | 76719 | Other injuries to scalp | Other injuries to scalp | nan | False |
| 17 | 810 | Murine (endemic) typhus | 812 | Scrub typhus; Fracture of humerus | Scrub typhus | Typhus | True | 802 | Intestinal infection due to enterotoxigenic E. coli; Fracture of face bones | Intestinal infection due to enterotoxigenic E. coli | nan | False |
| 18 | V281 | Antenatal screening for raised alpha-fetoprotein levels in amniotic fluid | V287 | | | nan | True | V270 | Outcome of delivery, single liveborn | Outcome of delivery, single liveborn | nan | False |

| | | | | | | | | | | | | |
|---|---|---|---|---|---|---|---|---|---|---|---|---|
| 19 | 30452 | Hallucinogen dependence, episodic | 3045 | Hallucinogen dependence | | nan | True | 3045 | Hallucinogen dependence | | nan | True |
| 20 | 64224 | Other pre-existing hypertension,complicating pregnancy, childbirth, and the puerperium, , postpartum condition or complication | 6427 | Pre-eclampsia or eclampsia superimposed on pre-existing hypertension | | nan | True | 6429 | Unspecified hypertension complicating pregnancy, childbirth, or the puerperium | | nan | True |
| 21 | 77182 | Urinary tract infection of newborn | 77182 | Urinary tract infection of newborn | Urinary tract infection of newborn | Exact Match | True | 77181 | Septicemia [sepsis] of newborn | Septicemia [sepsis] of newborn | Infection of newborn | True |
| 22 | 8977 | Traumatic amputation of leg(s) (complete) (partial), bilateral [any level], complicated | 8977 | Traumatic amputation of leg(s) (complete) (partial), bilateral [any level], complicated | Traumatic amputation of leg(s) (complete) (partial), bilateral [any level], complicated | Exact Match | True | 8959 | | | nan | False |
| 23 | 20531 | Myeloid sarcoma, in remission | 20501 | Acute myeloid leukemia, in remission | Acute myeloid leukemia, in remission | Myeloid cancer | True | 20871 | | | nan | False |
| 24 | 9134 | Insect bite, nonvenomous of elbow, forearm, and wrist, without mention of infection | 9194 | Insect bite, nonvenomous, of other, multiple, and unspecified sites, without mention of infection | Insect bite, nonvenomous, of other, multiple, and unspecified sites, without mention of infection | Insect bite | True | 9174 | Insect bite, nonvenomous, of foot and toe(s), without mention of infection | Insect bite, nonvenomous, of foot and toe(s), without mention of infection | Insect bite | True |
| 25 | 4838 | Pneumonia due to other specified organism | 486 | Pneumonia, organism unspecified | Pneumonia, organism unspecified | Pneumonia | True | 48289 | Pneumonia due to other specified bacteria | Pneumonia due to other specified bacteria | nan | True |
| 26 | 48882 | Influenza due to identified novel influenza A virus with other respiratory manifestations | 48811 | Influenza due to identified 2009 H1N1 influenza virus with pneumonia | Influenza due to identified 2009 H1N1 influenza virus with pneumonia | Influenza | False | 48819 | Influenza due to identified 2009 H1N1 influenza virus with other manifestations | Influenza due to identified 2009 H1N1 influenza virus with other manifestations | Influenza | False |
| 27 | V5872 | Aftercare following surgery of the nervous system, NEC | V5849 | Other specified aftercare following surgery | Other specified aftercare following surgery | Aftercare | False | V5881 | Fitting and adjustment of vascular catheter | Fitting and adjustment of vascular catheter | nan | False |
| 28 | 6220 | Erosion and ectropion of cervix | 6221 | Dysplasia of cervix (uteri) | | nan | True | 62211 | Mild dysplasia of cervix | Mild dysplasia of cervix | Cervical condition | True |
| 29 | 2707 | Other disturbances of straight-chain amino-acid metabolism | 2708 | Other specified disorders of amino-acid metabolism | Other specified disorders of amino-acid metabolism | Amino acid metabolism | True | 2708 | Other specified disorders of amino-acid metabolism | Other specified disorders of amino-acid metabolism | Amino acid metabolism | True |
| 30 | 78704 | Bilious emesis | 78703 | Vomiting alone | Vomiting alone | Vomiting | True | 577 | Diseases of pancreas | | nan | False |
| 31 | E9458 | Other and unspecified respiratory drugs causing adverse effects in therapeutic use | E9479 | Unspecified drug or medicinal substance causing adverse effects in therapeutic use | Unspecified drug or medicinal substance causing adverse effects in therapeutic use | Drug-related adverse event | False | 9952 | Other venereal diseases due to chlamydia trachomatis, anus and rectum; Other and unspecified adverse effect of drug, medicinal and biological substance | Other venereal diseases due to chlamydia trachomatis, anus and rectum | nan | False |
| 32 | 31220 | Socialized conduct disorder, unspecified | 3129 | Unspecified disturbance of conduct | Unspecified disturbance of conduct | Conduct disorder | True | 31281 | Conduct disorder, childhood onset type | Conduct disorder, childhood onset type | Conduct disorder | False |
| 33 | 1224 | Echinococcus granulosus infection, unspecified | 1229 | Echinococcosis, other and unspecified | Echinococcosis, other and unspecified | Echinococcus infection | True | 1229 | Echinococcosis, other and unspecified | Echinococcosis, other and unspecified | Echinococcus infection | True |
| 34 | 71126 | Arthropathy in Behcet's syndrome, lower leg | 7131 | Arthropathy associated with gastrointestinal conditions other than infections | Arthropathy associated with gastrointestinal conditions other than infections | Arthropathy related to gastrointestinal disease | False | 1361 | Tuberculous encephalitis or myelitis, bacteriological or histological examination not done; Behcet's syndrome | Tuberculous encephalitis or myelitis, bacteriological or histological examination not done; Behcet's syndrome | Behcet's syndrome | False |
| 35 | 30543 | Sedative, hypnotic or anxiolytic abuse, in remission | 3041 | Sedative, hypnotic or anxiolytic dependence | | nan | False | 30421 | Cocaine dependence, continuous | Cocaine dependence, continuous | Substance use | False |
| 36 | 74924 | Cleft palate with cleft lip, bilateral, incomplete | 7492 | Cleft palate with cleft lip | | nan | True | 7492 | Cleft palate with cleft lip | | nan | True |
| 37 | 76492 | Fetal growth retardation, unspecified, 500-749 grams | 764 | Slow fetal growth and fetal malnutrition | | nan | True | 765 | Disorders relating to short gestation and unspecified low birthweight | | nan | True |
| 38 | V2134 | Low birth weight status, 1500-1999 grams | 7641 | "Light-for-dates" with signs of fetal malnutrition | | nan | False | 76402 | "Light-for-dates" without mention of fetal malnutrition, 500-749 grams | "Light-for-dates" without mention of fetal malnutrition, 500-749 grams | Low birth weight | False |
| 39 | 308 | Other specified leprosy | 308 | Other specified leprosy; Acute reaction to stress | Other specified leprosy | Exact Match | True | 308 | Other specified leprosy; Acute reaction to stress | Other specified leprosy | Exact Match | True |

| | | | | | | | | | | | | |
|---|---|---|---|---|---|---|---|---|---|---|---|---|
| 40 | 80056 | Open fracture of vault of skull without mention of intracranial injury, with loss of consciousness of unspecified duration | 803 | Intestinal infection due to enteroinvasive E. coli; Other and unqualified skull fractures | | nan | True | 80002 | Closed fracture of vault of skull without mention of intracranial injury, with brief [less than one hour] loss of consciousness | Closed fracture of vault of skull without mention of intracranial injury, with brief [less than one hour] loss of consciousness | Skull fracture | False |
| 41 | 9882 | Toxic effect of berries and other plants eaten as food | E8631 | Accidental poisoning by insecticides of organophosphorus compounds | Accidental poisoning by insecticides of organophosphorus compounds | nan | False | 9892 | Toxic effect of chlorinated hydrocarbons | Toxic effect of chlorinated hydrocarbons | nan | True |
| 42 | 78832 | Stress incontinence, male | 78832 | Stress incontinence, male | Stress incontinence, male | Exact Match | True | 78832 | Stress incontinence, male | Stress incontinence, male | Exact Match | True |
| 43 | 5829 | Chronic glomerulonephritis with unspecified pathological lesion in kidney | 5829 | Other human herpesvirus encephalitis; Chronic glomerulonephritis with unspecified pathological lesion in kidney | Chronic glomerulonephritis with unspecified pathological lesion in kidney | Exact Match | True | 5839 | Nephritis and nephropathy, not specified as acute or chronic, with unspecified pathological lesion in kidney | Nephritis and nephropathy, not specified as acute or chronic, with unspecified pathological lesion in kidney | Nephritis | True |
| 44 | 30552 | Opioid abuse, episodic | 3055 | Opioid abuse | | nan | True | 30552 | Opioid abuse, episodic | Opioid abuse, episodic | Exact Match | True |
| 45 | V352 | Other multiple birth (three or more), mates all stillborn, born outside of hospital and not hospitalized | V273 | Outcome of delivery, twins, one liveborn and one stillborn | Outcome of delivery, twins, one liveborn and one stillborn | Multiple births with stillborn(s) | False | 65563 | Suspected damage to fetus from radiation, affecting management of mother, antepartum condition or complication | Suspected damage to fetus from radiation, affecting management of mother, antepartum condition or complication | Birth complication | False |
| 46 | 71622 | Allergic arthritis, upper arm | 71651 | Unspecified polyarthropathy or polyarthritis, shoulder region | Unspecified polyarthropathy or polyarthritis, shoulder region | Arthritis of upper arm | False | 71432 | Pauciarticular juvenile rheumatoid arthritis | Pauciarticular juvenile rheumatoid arthritis | Arthritis | False |
| 47 | 498 | Other specified non-arthropod-borne viral diseases of central nervous system | 498 | Other specified non-arthropod-borne viral diseases of central nervous system | Other specified non-arthropod-borne viral diseases of central nervous system | Exact Match | True | 3239 | Unspecified causes of encephalitis, myelitis, and encephalomyelitis | Unspecified causes of encephalitis, myelitis, and encephalomyelitis | nan | False |
| 48 | E8602 | Accidental poisoning by methyl alcohol | E8600 | Accidental poisoning by alcoholic beverages | Accidental poisoning by alcoholic beverages | Alcohol-based poisoning | True | 980 | Gonococcal infection (acute) of lower genitourinary tract; Toxic effect of alcohol | Gonococcal infection (acute) of lower genitourinary tract | nan | False |
| 49 | 6068 | Infertility due to extratesticular causes | 6069 | Male infertility, unspecified | Male infertility, unspecified | Male Infertility | True | 6069 | Male infertility, unspecified | Male infertility, unspecified | Male Infertility | True |
| 50 | 2702 | Other disturbances of aromatic amino-acid metabolism | 2702 | Other disturbances of aromatic amino-acid metabolism | Other disturbances of aromatic amino-acid metabolism | Exact Match | True | 2708 | Other specified disorders of amino-acid metabolism | Other specified disorders of amino-acid metabolism | Amino acid metabolism | True |
| 51 | 71537 | Osteoarthrosis, localized, not specified whether primary or secondary, ankle and foot | 71597 | Osteoarthrosis, unspecified whether generalized or localized, ankle and foot | Osteoarthrosis, unspecified whether generalized or localized, ankle and foot | Osteoarthrosis, ankle and foot | True | 71517 | Osteoarthrosis, localized, primary, ankle and foot | Osteoarthrosis, localized, primary, ankle and foot | Osteoarthrosis, ankle and foot | True |
| 52 | 34201 | Flaccid hemiplegia and hemiparesis affecting dominant side | 43821 | Late effects of cerebrovascular disease, hemiplegia affecting dominant side | Late effects of cerebrovascular disease, hemiplegia affecting dominant side | Hemiplegia, dominant side | False | 34201 | Flaccid hemiplegia and hemiparesis affecting dominant side | Flaccid hemiplegia and hemiparesis affecting dominant side | Exact Match | True |
| 53 | E8743 | Mechanical failure of instrument or apparatus during endoscopic examination | E8700 | Accidental cut, puncture, perforation or hemorrhage during surgical operation | Accidental cut, puncture, perforation or hemorrhage during surgical operation | Procedure complication | False | 99882 | Cataract fragments in eye following cataract surgery | Cataract fragments in eye following cataract surgery | Procedure complication | False |
| 54 | 3735 | Other infective dermatitis of eyelid | 3738 | Other inflammations of eyelids | Other inflammations of eyelids | Eyelid conditions | True | 37332 | Contact and allergic dermatitis of eyelid | Contact and allergic dermatitis of eyelid | Dermatitis of eyelid | True |
| 55 | 74904 | Cleft palate, bilateral, incomplete | 7491 | Cleft lip | | nan | True | 74924 | Cleft palate with cleft lip, bilateral, incomplete | Cleft palate with cleft lip, bilateral, incomplete | Cleft palate, bilateral, incomplete | True |
| 56 | E8143 | Motor vehicle traffic accident involving collision with pedestrian injuring passenger on motorcycle | E8127 | Other motor vehicle traffic accident involving collision with motor vehicle injuring pedestrian | Other motor vehicle traffic accident involving collision with motor vehicle injuring pedestrian | Motor vehicle accident involving pedestrian | False | E8126 | Other motor vehicle traffic accident involving collision with motor vehicle injuring pedal cyclist | Other motor vehicle traffic accident involving collision with motor vehicle injuring pedal cyclist | Motor vehicle accident involving individual outside of vehicle | False |
| 57 | 34120 | Acute (transverse) myelitis NOS | 3239 | Unspecified causes of encephalitis, myelitis, and encephalomyelitis | Unspecified causes of encephalitis, myelitis, and encephalomyelitis | Myelitis | False | 3239 | Unspecified causes of encephalitis, myelitis, and encephalomyelitis | Unspecified causes of encephalitis, myelitis, and encephalomyelitis | Myelitis | False |
| 58 | 38844 | Auditory recruitment | 38845 | Acquired auditory processing disorder | Acquired auditory processing disorder | Auditory disorder | True | 38842 | Hyperacusis | Hyperacusis | Auditory disorder | True |

| | | | | | | | | | | | | |
|---|---|---|---|---|---|---|---|---|---|---|---|---|
| 59 | 9987 | Acute reaction to foreign substance accidentally left during a procedure | 9987 | Acute reaction to foreign substance accidentally left during a procedure | Acute reaction to foreign substance accidentally left during a procedure | Exact Match | True | 9984 | Foreign body accidentally left during a procedure | Foreign body accidentally left during a procedure | Procedure complication | True |
| 60 | 417 | Pseudomonas infection in conditions classified elsewhere and of unspecified site | 417 | Pseudomonas infection in conditions classified elsewhere and of unspecified site; Other diseases of pulmonary circulation | Pseudomonas infection in conditions classified elsewhere and of unspecified site | Exact Match | True | 417 | Pseudomonas infection in conditions classified elsewhere and of unspecified site; Other diseases of pulmonary circulation | Pseudomonas infection in conditions classified elsewhere and of unspecified site | Exact Match | True |
| 61 | 7709 | Unspecified respiratory condition of fetus and newborn | 7709 | Unspecified respiratory condition of fetus and newborn | Unspecified respiratory condition of fetus and newborn | Exact Match | True | 7708 | Other newborn respiratory problems | | nan | True |
| 62 | 9511 | Injury to trochlear nerve | 9515 | Injury to acoustic nerve | Injury to acoustic nerve | Nerve injuries | True | 37681 | Orbital cysts | Orbital cysts | nan | False |
| 63 | V1240 | Personal history of unspecified disorder of nervous system and sense organs | V1259 | Personal history of other diseases of circulatory system | Personal history of other diseases of circulatory system | nan | False | V158 | Other specified personal history presenting hazards to health | | nan | False |
| 64 | 70713 | Ulcer of ankle | 70715 | Ulcer of other part of foot | Ulcer of other part of foot | Ankle or foot ulcer | True | 70712 | Ulcer of calf | Ulcer of calf | Lower limb ulcer | True |
| 65 | 2532 | Panhypopituitarism | 2532 | Panhypopituitarism | Panhypopituitarism | Exact Match | True | 2534 | Other anterior pituitary disorders | Other anterior pituitary disorders | Pituitary disorder | True |
| 66 | 7104 | Polymyositis | 7104 | Polymyositis | Polymyositis | Exact Match | True | 7104 | Polymyositis | Polymyositis | Exact Match | True |
| 67 | 42789 | Other specified cardiac dysrhythmias | 42789 | Other specified cardiac dysrhythmias | Other specified cardiac dysrhythmias | Exact Match | True | 42789 | Other specified cardiac dysrhythmias | Other specified cardiac dysrhythmias | Exact Match | True |
| 68 | 48230 | Pneumonia due to Streptococcus, unspecified | 481 | Pneumococcal pneumonia [Streptococcus pneumoniae pneumonia] | Pneumococcal pneumonia [Streptococcus pneumoniae pneumonia] | Streptococcal Pneumonia | True | 481 | Pneumococcal pneumonia [Streptococcus pneumoniae pneumonia] | Pneumococcal pneumonia [Streptococcus pneumoniae pneumonia] | Streptococcal Pneumonia | True |
| 69 | 36061 | Foreign body in anterior chamber | 36063 | Foreign body in lens | Foreign body in lens | Foreign body in eye | True | 36021 | Progressive high (degenerative) myopia | Progressive high (degenerative) myopia | Eye disorder | True |
| 70 | V6282 | Bereavement, uncomplicated | V6282 | Bereavement, uncomplicated | Bereavement, uncomplicated | Exact Match | True | V6282 | Bereavement, uncomplicated | Bereavement, uncomplicated | Exact Match | True |
| 71 | V5872 | Aftercare following surgery of the nervous system, NEC | V5872 | Aftercare following surgery of the nervous system, NEC | Aftercare following surgery of the nervous system, NEC | Exact Match | True | V5889 | Other specified aftercare | Other specified aftercare | Aftercare following surgery | False |
| 72 | 52340 | Chronic periodontitis, unspecified | 5234 | Chronic periodontitis | | nan | True | 523 | Gingival and periodontal diseases | | nan | True |
| 73 | 9852 | Toxic effect of manganese and its compounds | 985 | Gonococcal infection of joint; Toxic effect of other metals | | nan | True | 9851 | Gonococcal synovitis and tenosynovitis; Toxic effect of arsenic and its compounds | Toxic effect of arsenic and its compounds | Metal toxicosis | True |
| 74 | V1324 | Personal history of vulvar dysplasia | V1329 | Personal history of other genital system and obstetric disorders | Personal history of other genital system and obstetric disorders | Personal history of genital system disorder | True | V1389 | Personal history of other specified diseases | Personal history of other specified diseases | nan | False |
| 75 | 73082 | Other infections involving bone in diseases classified elsewhere, upper arm | 73021 | Unspecified osteomyelitis, shoulder region | Unspecified osteomyelitis, shoulder region | Bone infections, upper limb/shoulder | False | 73028 | Unspecified osteomyelitis, other specified sites | Unspecified osteomyelitis, other specified sites | Bone infections | False |
| 76 | 1902 | Malignant neoplasm of lacrimal gland | 1906 | Malignant neoplasm of choroid | Malignant neoplasm of choroid | Malignant neoplasm of eye | True | 193 | Malignant neoplasm of thyroid gland | Malignant neoplasm of thyroid gland | nan | True |
| 77 | 27401 | Acute gouty arthropathy | 27401 | Acute gouty arthropathy | Acute gouty arthropathy | Exact Match | True | 274 | Gout | | nan | True |
| 78 | 2922 | Pathological drug intoxication | 2929 | Unspecified drug-induced mental disorder | Unspecified drug-induced mental disorder | Drug-related change in mental status | True | 2929 | Unspecified drug-induced mental disorder | Unspecified drug-induced mental disorder | Drug-related change in mental status | True |
| 79 | 76514 | Other preterm infants, 1,000-1,249 grams | 76524 | 27-28 completed weeks of gestation | 27-28 completed weeks of gestation | Premature birth | True | V3012 | | | nan | False |
| 80 | 96900 | Poisoning by antidepressant, unspecified | 969 | Poisoning by psychotropic agents | | nan | True | 9699 | Poisoning by unspecified psychotropic agent | Poisoning by unspecified psychotropic agent | Psychotropic-induced poisoning | True |
| 81 | 9171 | Abrasion or friction burn of foot and toe(s), infected | 9447 | | | nan | False | 9178 | Other and unspecified superficial injury of foot and toes, without mention of infection | Other and unspecified superficial injury of foot and toes, without mention of infection | Foot and toe injuries | True |
| 82 | 94407 | Burn of unspecified degree of wrist | 94427 | Blisters, epidermal loss [second degree] of wrist | Blisters, epidermal loss [second degree] of wrist | Wrist burn injury | True | 9412 | Blisters with epidermal loss due to burn [second degree] of face, head, and neck | | nan | False |
| 83 | 7675 | Facial nerve injury due to birth trauma | 7676 | Injury to brachial plexus due to birth trauma | Injury to brachial plexus due to birth trauma | Birth trauma injury | True | 7674 | Injury to spine and spinal cord due to birth trauma | Injury to spine and spinal cord due to birth trauma | Nerve injury due to birth trauma | True |

| | | | | | | | | | | | | |
|---|---|---|---|---|---|---|---|---|---|---|---|---|
| 84 | 69514 | Stevens-Johnson syndrome-toxic epidermal necrolysis overlap syndrome | 69515 | Toxic epidermal necrolysis | Toxic epidermal necrolysis | Toxic epidermal necrolysis | True | 69514 | Stevens-Johnson syndrome-toxic epidermal necrolysis overlap syndrome | Stevens-Johnson syndrome-toxic epidermal necrolysis overlap syndrome | Exact Match | True |
| 85 | 86610 | Injury to kidney with open wound into cavity, unspecified injury | 866 | Enteritis due to astrovirus; Injury to kidney | | nan | True | 866 | Enteritis due to astrovirus; Injury to kidney | | nan | True |
| 86 | 1322 | Tuberculoma of brain, bacteriological or histological examination unknown (at present) | 13 | Tuberculosis of meninges and central nervous system | | nan | True | 13 | Tuberculosis of meninges and central nervous system | | nan | True |
| 87 | E9914 | Injury due to war operations by fragments from munitions | E9981 | Injury due to war operations but occurring after cessation of hostilities by explosion of bombs | Injury due to war operations but occurring after cessation of hostilities by explosion of bombs | War-related injury | False | E9880 | Injury by jumping or lying before moving object, undetermined whether accidentally or purposely inflicted | Injury by jumping or lying before moving object, undetermined whether accidentally or purposely inflicted | nan | False |
| 88 | E9492 | Typhus vaccine causing adverse effects in therapeutic use | 99529 | Unspecified adverse effect of other drug, medicinal and biological substance | Unspecified adverse effect of other drug, medicinal and biological substance | Adverse drug event | False | 99949 | Anaphylactic reaction due to other serum | Anaphylactic reaction due to other serum | Adverse drug event | False |
| 89 | 20916 | Malignant carcinoid tumor of the sigmoid colon | 20925 | Malignant carcinoid tumor of foregut, not otherwise specified | Malignant carcinoid tumor of foregut, not otherwise specified | Malignant gastrointestinal carcinoid tumor | False | 20952 | Benign carcinoid tumor of the cecum | Benign carcinoid tumor of the cecum | Gastrointestinal carcinoid tumor | False |
| 90 | V5971 | Egg (oocyte) (ovum) donor, under age 35, anonymous recipient | V2631 | Testing of female for genetic disease carrier status | Testing of female for genetic disease carrier status | nan | False | V2621 | Fertility testing | Fertility testing | nan | False |
| 91 | 8870 | Traumatic amputation of arm and hand (complete) (partial), unilateral, below elbow, without mention of complication | 887 | Traumatic amputation of arm and hand (complete) (partial) | | nan | True | 887 | Traumatic amputation of arm and hand (complete) (partial) | | nan | True |
| 92 | 5644 | Other postoperative functional disorders | 9979 | Complications affecting other specified body systems, not elsewhere classified | | nan | False | 99739 | Other respiratory complications | Other respiratory complications | Postoperative complications | False |
| 93 | 23989 | Neoplasms of unspecified nature, other specified sites | 23989 | Neoplasms of unspecified nature, other specified sites | Neoplasms of unspecified nature, other specified sites | Exact Match | True | 2399 | Neoplasm of unspecified nature, site unspecified | Neoplasm of unspecified nature, site unspecified | Unspecified neoplasms | True |
| 94 | 1005 | Primary tuberculous infection, tubercle bacilli not found by bacteriological examination, but tuberculosis confirmed histologically | 1085 | Other primary progressive tuberculosis, tubercle bacilli not found by bacteriological examination, but tuberculosis confirmed histologically | Other primary progressive tuberculosis, tubercle bacilli not found by bacteriological examination, but tuberculosis confirmed histologically | Primary tuberculosis | True | 101 | Tuberculous pleurisy in primary progressive tuberculosis; Vincent's angina | Vincent's angina | nan | True |
| 95 | 1655 | Tuberculosis of other male genital organs, tubercle bacilli not found by bacteriological examination, but tuberculosis confirmed histologically | 169 | Genitourinary tuberculosis, unspecified | | nan | True | 167 | Tuberculosis of other female genital organs | | nan | True |
| 96 | 5739 | Unspecified disorder of liver | 5739 | Unspecified disorder of liver | Unspecified disorder of liver | Exact Match | True | 5739 | Unspecified disorder of liver | Unspecified disorder of liver | Exact Match | True |
| 97 | 9771 | Poisoning by lipotropic drugs | 9778 | Poisoning by other specified drugs and medicinal substances | Poisoning by other specified drugs and medicinal substances | Drug poisoning | True | 9655 | Poisoning by pyrazole derivatives | Poisoning by pyrazole derivatives | nan | False |
| 98 | 6823 | Cellulitis and abscess of upper arm and forearm | 6823 | Cellulitis and abscess of upper arm and forearm | Cellulitis and abscess of upper arm and forearm | Exact Match | True | 6824 | Cellulitis and abscess of hand, except fingers and thumb | Cellulitis and abscess of hand, except fingers and thumb | nan | True |
| 99 | 94412 | Erythema [first degree] of thumb (nail) | 94402 | Burn of unspecified degree of thumb (nail) | Burn of unspecified degree of thumb (nail) | Thumb burn injury | True | 692 | Contact dermatitis and other eczema | | nan | False |
| 100 | 30722 | Chronic motor or vocal tic disorder | 30723 | Tourette's disorder | Tourette's disorder | Tic disorder | True | 30721 | Transient tic disorder | Transient tic disorder | Tic disorders | True |

Supplementary Table 2: Code-level generated outputs and similarity for ICD-10-CM

| Case | Original ICD-10-CM code | Original ICD-10-CM code description | GPT-4 Code | Description matching GPT-4 code | Description matching billable GPT-4 code | GPT-4 code semantic similarity | GPT-4 code syntactic similarity | GPT-3.5 Code | Description matching GPT-3.5 code | Description matching billable GPT-3.5 code | GPT-3.5 code semantic similarity | GPT-3.5 code syntactic similarity |
|---|---|---|---|---|---|---|---|---|---|---|---|---|
| 1 | S82433B | Displaced oblique fracture of shaft of unspecified fibula, initial encounter for open fracture type I or II | S82401B | Unspecified fracture of shaft of right fibula, initial encounter for open fracture type I or II | Unspecified fracture of shaft of right fibula, initial encounter for open fracture type I or II | Fibula fracture, initial encounter | False | S82424A | Nondisplaced transverse fracture of shaft of right fibula, initial encounter for closed fracture | Nondisplaced transverse fracture of shaft of right fibula, initial encounter for closed fracture | Fibula fracture, initial encounter | False |
| 2 | S82035B | Nondisplaced transverse fracture of left patella, initial encounter for open fracture type I or II | S82112B | Displaced fracture of left tibial spine, initial encounter for open fracture type I or II | Displaced fracture of left tibial spine, initial encounter for open fracture type I or II | Fracture of knee joint bones, initial encounter | False | S82102A | Unspecified fracture of upper end of left tibia, initial encounter for closed fracture | Unspecified fracture of upper end of left tibia, initial encounter for closed fracture | Fracture of knee joint bones, initial encounter | False |
| 3 | S72444P | Nondisplaced fracture of lower epiphysis (separation) of right femur, subsequent encounter for closed fracture with malunion | S72353K | Displaced comminuted fracture of shaft of unspecified femur, subsequent encounter for closed fracture with nonunion | Displaced comminuted fracture of shaft of unspecified femur, subsequent encounter for closed fracture with nonunion | Femur fracture, subsequent encounter | False | S72113S | Displaced fracture of greater trochanter of unspecified femur, sequela | Displaced fracture of greater trochanter of unspecified femur, sequela | Femur fracture | False |
| 4 | M8450XP | Pathological fracture in neoplastic disease, unspecified site, subsequent encounter for fracture with malunion | M8450K | | | nan | True | M8440XS | Pathological fracture, unspecified site, sequela | Pathological fracture, unspecified site, sequela | Pathological fracture | False |
| 5 | S62301D | Unspecified fracture of second metacarpal bone, left hand, subsequent encounter for fracture with routine healing | S62522D | Displaced fracture of distal phalanx of left thumb, subsequent encounter for fracture with routine healing | Displaced fracture of distal phalanx of left thumb, subsequent encounter for fracture with routine healing | Left hand fracture with routine healing, subsequent encounter | False | S62352S | Nondisplaced fracture of shaft of third metacarpal bone, right hand, sequela | Nondisplaced fracture of shaft of third metacarpal bone, right hand, sequela | Metacarpal fracture with routine healing | False |
| 6 | S72362N | Displaced segmental fracture of shaft of left femur, subsequent encounter for open fracture type IIIA, IIIB, or IIIC with nonunion | S72352M | Displaced comminuted fracture of shaft of left femur, subsequent encounter for open fracture type I or II with nonunion | Displaced comminuted fracture of shaft of left femur, subsequent encounter for open fracture type I or II with nonunion | Open fracture of the left femur with nonunion, subsequent encounter | False | S72342K | Displaced spiral fracture of shaft of left femur, subsequent encounter for closed fracture with nonunion | Displaced spiral fracture of shaft of left femur, subsequent encounter for closed fracture with nonunion | Fracture of the left femur with nonunion, subsequent encounter | False |
| 7 | I82401 | Acute embolism and thrombosis of unspecified deep veins of right lower extremity | I82409 | Acute embolism and thrombosis of unspecified deep veins of unspecified lower extremity | Acute embolism and thrombosis of unspecified deep veins of unspecified lower extremity | Acute embolism and thrombosis of deep veins of lower extremity | True | I82609 | Acute embolism and thrombosis of unspecified veins of unspecified upper extremity | Acute embolism and thrombosis of unspecified veins of unspecified upper extremity | Acute embolism and thrombosis of deep veins | False |
| 8 | S52552F | Other extraarticular fracture of lower end of left radius, subsequent encounter for open fracture type IIIA, IIIB, or IIIC with routine healing | S52526D | | | nan | False | S52309D | Unspecified fracture of shaft of unspecified radius, subsequent encounter for closed fracture with routine healing | Unspecified fracture of shaft of unspecified radius, subsequent encounter for closed fracture with routine healing | Radius fracture with routine healing, subsequent encounter | False |
| 9 | S86391S | Other injury of muscle(s) and tendon(s) of peroneal muscle group at lower leg level, right leg, sequela | S86891S | Other injury of other muscle(s) and tendon(s) at lower leg level, right leg, sequela | Other injury of other muscle(s) and tendon(s) at lower leg level, right leg, sequela | Other injury of other muscle(s) and tendon(s) at lower leg level, right leg, sequela | True | S86829S | Laceration of other muscle(s) and tendon(s) at lower leg level, unspecified leg, sequela | Laceration of other muscle(s) and tendon(s) at lower leg level, unspecified leg, sequela | Injury of other muscle(s) and tendon(s) at lower leg level, sequela | False |
| 10 | V8082XS | Occupant of animal-drawn vehicle injured in collision with fixed or stationary object, sequela | V80719S | | | nan | False | V819XXS | Occupant of railway train or railway vehicle injured in unspecified railway accident, sequela | Occupant of railway train or railway vehicle injured in unspecified railway accident, sequela | Injury related to vehicular accident | False |
| 11 | S96992S | Other specified injury of unspecified muscle and tendon at ankle and foot level, left foot, sequela | S96892S | Other specified injury of other specified muscles and tendons at ankle and foot level, left foot, sequela | Other specified injury of other specified muscles and tendons at ankle and foot level, left foot, sequela | Other specified injury of muscle and tendon at ankle and foot level, left foot, sequela | True | S99892S | | | nan | False |
| 12 | S20461A | Insect bite (nonvenomous) of right back wall of thorax, initial encounter | S20319A | Abrasion of unspecified front wall of thorax, initial encounter | Abrasion of unspecified front wall of thorax, initial encounter | nan | False | S30824A | Blister (nonthermal) of vagina and vulva, initial encounter | Blister (nonthermal) of vagina and vulva, initial encounter | nan | False |
| 13 | M5116 | Intervertebral disc disorders with radiculopathy, lumbar region | M5116 | Intervertebral disc disorders with radiculopathy, lumbar region | Intervertebral disc disorders with radiculopathy, lumbar region | Exact Match | True | M5116 | Intervertebral disc disorders with radiculopathy, lumbar region | Intervertebral disc disorders with radiculopathy, lumbar region | Exact Match | True |

| | | | | | | | | | | | | |
|---|---|---|---|---|---|---|---|---|---|---|---|---|
| 14 | Y93B1 | Activity, exercise machines primarily for muscle strengthening | Y9364 | Activity, softball | Activity, baseball | Physical activities | False | Z742 | Need for assistance at home and no other household member able to render care | Need for assistance at home and no other household member able to render care | nan | False |
| 15 | D3A023 | Benign carcinoid tumor of the transverse colon | D3A020 | Benign carcinoid tumor of the appendix | Benign carcinoid tumor of the appendix | Benign carcinoid tumors of lower gastrointestinal tract | True | D3A00 | Benign carcinoid tumor of unspecified site | Benign carcinoid tumor of unspecified site | Benign carcinoid tumor | True |
| 16 | T462X6S | Underdosing of other antidysrhythmic drugs, sequela | T461X5S | Adverse effect of calcium-channel blockers, sequela | Adverse effect of calcium-channel blockers, sequela | nan | False | T468X5S | Adverse effect of antivaricose drugs, including sclerosing agents, sequela | Adverse effect of antivaricose drugs, including sclerosing agents, sequela | nan | False |
| 17 | T398X3A | Poisoning by other nonopioid analgesics and antipyretics, not elsewhere classified, assault, initial encounter | T39095A | Adverse effect of salicylates, initial encounter | Adverse effect of salicylates, initial encounter | Anelgesic-related adverse event, initial encounter | False | X424XXA | | | nan | False |
| 18 | S52102S | Unspecified fracture of upper end of left radius, sequela | S52502S | Unspecified fracture of the lower end of left radius, sequela | Unspecified fracture of the lower end of left radius, sequela | Unspecified fracture of the left radius, sequela | True | S5232XS | | | nan | False |
| 19 | T63043A | Toxic effect of cobra venom, assault, initial encounter | T630X2A | | | nan | False | T63121A | Toxic effect of venom of other venomous lizard, accidental (unintentional), initial encounter | Toxic effect of venom of other venomous lizard, accidental (unintentional), initial encounter | Toxic effect of venom, initial encounter | False |
| 20 | W5913XA | Crushed by nonvenomous snake, initial encounter | W5922XA | Struck by turtle, initial encounter | Struck by turtle, initial encounter | Mechanical injury by animal, initial encounter | False | W5912XA | Struck by nonvenomous snake, initial encounter | Struck by nonvenomous snake, initial encounter | Mechanical injury by snake, initial encounter | True |
| 21 | C969 | Malignant neoplasm of lymphoid, hematopoietic and related tissue, unspecified | C859 | Lymphoma NOS | | nan | False | C969 | Malignant neoplasm of lymphoid, hematopoietic and related tissue, unspecified | Malignant neoplasm of lymphoid, hematopoietic and related tissue, unspecified | Exact Match | True |
| 22 | S62234B | Other nondisplaced fracture of base of first metacarpal bone, right hand, initial encounter for open fracture | S62241B | Displaced fracture of shaft of first metacarpal bone, right hand, initial encounter for open fracture | Displaced fracture of shaft of first metacarpal bone, right hand, initial encounter for open fracture | Fracture of the first metacarpal bone in the right hand, initial encounter for open fracture | False | S62414A | | | nan | False |
| 23 | S63427A | Traumatic rupture of palmar ligament of left little finger at metacarpophalangeal and interphalangeal joint, initial encounter | S63625A | | | nan | False | S63664A | | | nan | False |
| 24 | V5910XA | Passenger in pick-up truck or van injured in collision with unspecified motor vehicles in nontraffic accident, initial encounter | V877XXA | Person injured in collision between other specified motor vehicles (traffic), initial encounter | Person injured in collision between other specified motor vehicles (traffic), initial encounter | Person injury in collision between motor vehicles, intial encounter | False | V579XXA | Unspecified occupant of pick-up truck or van injured in collision with fixed or stationary object in traffic accident, initial encounter | Unspecified occupant of pick-up truck or van injured in collision with fixed or stationary object in traffic accident, initial encounter | Occupant of pick-up truck or van injured in collision between motor vehicles, intial encounter | False |
| 25 | W9423XA | Exposure to sudden change in air pressure in aircraft during ascent, initial encounter | W941XXA | | | nan | False | W940 | Exposure to prolonged high air pressure | | nan | True |
| 26 | D030 | Melanoma in situ of lip | D030 | Melanoma in situ of lip | Melanoma in situ of lip | Exact Match | True | D0361 | Melanoma in situ of right upper limb, including shoulder | Melanoma in situ of right upper limb, including shoulder | Melanoma in situ | True |
| 27 | H01023 | Squamous blepharitis right eye, unspecified eyelid | H01003 | Unspecified blepharitis right eye, unspecified eyelid | Unspecified blepharitis right eye, unspecified eyelid | Blepharitis right eye, unspecified eyelid | True | H01139 | Eczematous dermatitis of unspecified eye, unspecified eyelid | Eczematous dermatitis of unspecified eye, unspecified eyelid | Eyelid conditions | False |
| 28 | V2139XS | Person boarding or alighting other motorcycle injured in collision with pedal cycle, sequela | V20421S | | | nan | False | V1940XA | Pedal cycle driver injured in collision with unspecified motor vehicles in traffic accident, initial encounter | Pedal cycle driver injured in collision with unspecified motor vehicles in traffic accident, initial encounter | Injury after collision of pedal cycle driver with vehicle | False |
| 29 | S72353C | Displaced comminuted fracture of shaft of unspecified femur, initial encounter for open fracture type IIIA, IIIB, or IIIC | S72901B | | | nan | False | S72301A | Unspecified fracture of shaft of right femur, initial encounter for closed fracture | Unspecified fracture of shaft of right femur, initial encounter for closed fracture | Femur fracture, initial encounter | False |

| | | | | | | | | | | | | |
|---|---|---|---|---|---|---|---|---|---|---|---|---|
| 30 | S75102S | Unspecified injury of femoral vein at hip and thigh level, left leg, sequela | S75892S | Other specified injury of other blood vessels at hip and thigh level, left leg, sequela | Other specified injury of other blood vessels at hip and thigh level, left leg, sequela | Injury to blood vessel at hip and thigh level, left leg, sequela | False | S35042S | | | nan | False |
| 31 | S41102A | Unspecified open wound of left upper arm, initial encounter | S41102A | Unspecified open wound of left upper arm, initial encounter | Unspecified open wound of left upper arm, initial encounter | Exact Match | True | S51111A | | | nan | False |
| 32 | S022XXB | Fracture of nasal bones, initial encounter for open fracture | S022XXB | Fracture of nasal bones, initial encounter for open fracture | Fracture of nasal bones, initial encounter for open fracture | Exact Match | True | S021XXA | | | nan | False |
| 33 | M6289 | Other specified disorders of muscle | M6289 | Muscle (sheath) hernia | Other specified disorders of muscle | Exact Match | True | M6289 | Muscle (sheath) hernia | Other specified disorders of muscle | Exact Match | True |
| 34 | S52334A | Nondisplaced oblique fracture of shaft of right radius, initial encounter for closed fracture | S52321A | Displaced transverse fracture of shaft of right radius, initial encounter for closed fracture | Displaced transverse fracture of shaft of right radius, initial encounter for closed fracture | Closed fracture of the shaft of the right radius, initial encounter | False | S52212A | Greenstick fracture of shaft of left ulna, initial encounter for closed fracture | Greenstick fracture of shaft of left ulna, initial encounter for closed fracture | Closed fracture of the shaft of the lower arm, initial encounter | False |
| 35 | Q163 | Congenital malformation of ear ossicles | Q163 | Congenital fusion of ear ossicles | Congenital malformation of ear ossicles | Exact Match | True | Q160 | Congenital absence of (ear) auricle | Congenital absence of (ear) auricle | Ear malformation | True |
| 36 | S52225H | Nondisplaced transverse fracture of shaft of left ulna, subsequent encounter for open fracture type I or II with delayed healing | S52222D | Displaced transverse fracture of shaft of left ulna, subsequent encounter for closed fracture with routine healing | Displaced transverse fracture of shaft of left ulna, subsequent encounter for closed fracture with routine healing | Transverse fracture of the shaft of the left ulna, subsequent encounter | False | S52212K | Greenstick fracture of shaft of left ulna, subsequent encounter for fracture with nonunion | Greenstick fracture of shaft of left ulna, subsequent encounter for fracture with nonunion | Fracture of the shaft of the left ulna, subsequent encounter | False |
| 37 | Q175 | Prominent ear | H61109 | Unspecified noninfective disorders of pinna, unspecified ear | Unspecified noninfective disorders of pinna, unspecified ear | Ear condition | False | Q173 | Pointed ear | Other misshapen ear | Ear deformities | True |
| 38 | S86819S | Strain of other muscle(s) and tendon(s) at lower leg level, unspecified leg, sequela | S86919S | Strain of unspecified muscle(s) and tendon(s) at lower leg level, unspecified leg, sequela | Strain of unspecified muscle(s) and tendon(s) at lower leg level, unspecified leg, sequela | Strain of muscles and tendons at lower leg level, unspecified leg, sequela | True | S86919S | Strain of unspecified muscle(s) and tendon(s) at lower leg level, unspecified leg, sequela | Strain of unspecified muscle(s) and tendon(s) at lower leg level, unspecified leg, sequela | Strain of muscles and tendons at lower leg level, unspecified leg, sequela | True |
| 39 | S82152A | Displaced fracture of left tibial tuberosity, initial encounter for closed fracture | S82122A | Displaced fracture of lateral condyle of left tibia, initial encounter for closed fracture | Displaced fracture of lateral condyle of left tibia, initial encounter for closed fracture | Displaced fracture of lateral left tibia, initial encounter for closed fracture | True | S82202A | Unspecified fracture of shaft of left tibia, initial encounter for closed fracture | Unspecified fracture of shaft of left tibia, initial encounter for closed fracture | Displaced fracture of left tibia, initial encounter for closed fracture | False |
| 40 | H4010X4 | Unspecified open-angle glaucoma, indeterminate stage | H4011X0 | | | nan | False | H4010X0 | Unspecified open-angle glaucoma, stage unspecified | Unspecified open-angle glaucoma, stage unspecified | Unspecified open-angle glaucoma, unclear stage | True |
| 41 | T480X1S | Poisoning by oxytocic drugs, accidental (unintentional), sequela | T384X1S | Poisoning by oral contraceptives, accidental (unintentional), sequela | Poisoning by oral contraceptives, accidental (unintentional), sequela | Poisoning by hormonal drugs | False | T485X6S | Underdosing of other anti-common-cold drugs, sequela | Underdosing of other anti-common-cold drugs, sequela | nan | False |
| 42 | D483 | Neoplasm of uncertain behavior of retroperitoneum | D483 | Neoplasm of uncertain behavior of retroperitoneum | Neoplasm of uncertain behavior of retroperitoneum | Exact Match | True | D481 | Neoplasm of uncertain behavior of connective and other soft tissue | Neoplasm of uncertain behavior of connective and other soft tissue | Neoplasm of uncertain behavior | True |
| 43 | T24609D | Corrosion of second degree of unspecified site of unspecified lower limb, except ankle and foot, subsequent encounter | T24909D | | | nan | True | T24462S | | | nan | False |
| 44 | T50B96S | Underdosing of other viral vaccines, sequela | T50B95S | Adverse effect of other viral vaccines, sequela | Adverse effect of other viral vaccines, sequela | Issues with viral vaccines, sequela | True | T8803XS | | | nan | False |
| 45 | Q984 | Klinefelter syndrome, unspecified | Q989 | Sex chromosome abnormality, male phenotype, unspecified | Sex chromosome abnormality, male phenotype, unspecified | Male sex chromosome abnormalities | True | Q980 | Klinefelter syndrome karyotype 47, XXY | Klinefelter syndrome karyotype 47, XXY | Klinefelter syndrome | True |
| 46 | E441 | Mild protein-calorie malnutrition | E440 | Moderate protein-calorie malnutrition | Moderate protein-calorie malnutrition | Protein-calorie malnutrition | True | E43 | Unspecified severe protein-calorie malnutrition | Unspecified severe protein-calorie malnutrition | Protein-calorie malnutrition | True |
| 47 | S61232D | Puncture wound without foreign body of right middle finger without damage to nail, subsequent encounter | S61432D | Puncture wound without foreign body of left hand, subsequent encounter | Puncture wound without foreign body of left hand, subsequent encounter | Puncture wound without foreign body of hand, subsequent encounter | True | S61441D | Puncture wound with foreign body of right hand, subsequent encounter | Puncture wound with foreign body of right hand, subsequent encounter | Puncture wound of right hand, subsequent encounter | False |
| 48 | M23005 | Cystic meniscus, unspecified medial meniscus, unspecified knee | M23859 | | | nan | False | M2320 | Derangement of unspecified lateral meniscus due to old tear or injury | | nan | True |

| | | | | | | | | | | | | |
|---|---|---|---|---|---|---|---|---|---|---|---|---|
| 49 | S06365D | Traumatic hemorrhage of cerebrum, unspecified, with loss of consciousness greater than 24 hours with return to pre-existing conscious level, subsequent encounter | S069X0S | Unspecified intracranial injury without loss of consciousness, sequela | Unspecified intracranial injury without loss of consciousness, sequela | Unspecified intracranial injury without loss of consciousness | False | S068X9S | | | nan | False |
| 50 | S72101H | Unspecified trochanteric fracture of right femur, subsequent encounter for open fracture type I or II with delayed healing | S7214XD | | | nan | False | S72145S | Nondisplaced intertrochanteric fracture of left femur, sequela | Nondisplaced intertrochanteric fracture of left femur, sequela | Femur fracture | False |
| 51 | A403 | Sepsis due to Streptococcus pneumoniae | A403 | Pneumococcal sepsis | Sepsis due to Streptococcus pneumoniae | Exact Match | True | A403 | Pneumococcal sepsis | Sepsis due to Streptococcus pneumoniae | Exact Match | True |
| 52 | T23569D | Corrosion of first degree of back of unspecified hand, subsequent encounter | T25011D | Burn of unspecified degree of right ankle, subsequent encounter | Burn of unspecified degree of right ankle, subsequent encounter | Burn injury, subsequent encounter | False | T23231S | Burn of second degree of multiple right fingers (nail), not including thumb, sequela | Burn of second degree of multiple right fingers (nail), not including thumb, sequela | Burn injury of hand | False |
| 53 | T82321A | Displacement of carotid arterial graft (bypass), initial encounter | T82858A | Stenosis of other vascular prosthetic devices, implants and grafts, initial encounter | Stenosis of other vascular prosthetic devices, implants and grafts, initial encounter | Vascular graft complication, initial encounter | False | T82858A | Stenosis of other vascular prosthetic devices, implants and grafts, initial encounter | Stenosis of other vascular prosthetic devices, implants and grafts, initial encounter | Vascular graft complication, initial encounter | False |
| 54 | S42424A | Nondisplaced comminuted supracondylar fracture without intercondylar fracture of right humerus, initial encounter for closed fracture | S42454A | Nondisplaced fracture of lateral condyle of right humerus, initial encounter for closed fracture | Nondisplaced fracture of lateral condyle of right humerus, initial encounter for closed fracture | Nondisplaced fracture of right humerus, initial encounter for closed fracture | True | S42421A | Displaced comminuted supracondylar fracture without intercondylar fracture of right humerus, initial encounter for closed fracture | Displaced comminuted supracondylar fracture without intercondylar fracture of right humerus, initial encounter for closed fracture | Comminuted supracondylar fracture without intercondylar fracture of right humerus, initial encounter for closed fracture | True |
| 55 | O3513X9 | Maternal care for (suspected) chromosomal abnormality in fetus, Trisomy 21, other fetus | O352X2 | | | nan | False | O357XX0 | Maternal care for (suspected) damage to fetus by other medical procedures, not applicable or unspecified | Maternal care for (suspected) damage to fetus by other medical procedures, not applicable or unspecified | Maternal care for suspected fetal condition | False |
| 56 | H59323 | Postprocedural hemorrhage of eye and adnexa following other procedure, bilateral | H59021 | Cataract (lens) fragments in eye following cataract surgery, right eye | Cataract (lens) fragments in eye following cataract surgery, right eye | Postprocedural complications in the eye | False | H59023 | Cataract (lens) fragments in eye following cataract surgery, bilateral | Cataract (lens) fragments in eye following cataract surgery, bilateral | Postprocedural complications in bilateral eyes | True |
| 57 | V9137XA | Hit or struck by falling object due to accident to water-skis, initial encounter | V9107XA | Burn due to water-skis on fire, initial encounter | Burn due to water-skis on fire, initial encounter | Injury related to water-skis, initial encounter | True | V9107XA | Burn due to water-skis on fire, initial encounter | Burn due to water-skis on fire, initial encounter | Injury related to water-skis, initial encounter | True |
| 58 | W5522XD | Struck by cow, subsequent encounter | W5521XD | Bitten by cow, subsequent encounter | Bitten by cow, subsequent encounter | Injury related to cow, subsequent encounter | True | W5521XS | Bitten by cow, sequela | Bitten by cow, sequela | Injury related to cow, subsequent encounter | False |
| 59 | S71141A | Puncture wound with foreign body, right thigh, initial encounter | S71141A | Puncture wound with foreign body, right thigh, initial encounter | Puncture wound with foreign body, right thigh, initial encounter | Exact Match | True | S81821A | Laceration with foreign body, right lower leg, initial encounter | Laceration with foreign body, right lower leg, initial encounter | Right lower leg injury with foreign body, initial encounter | False |
| 60 | E083543 | Diabetes mellitus due to underlying condition with proliferative diabetic retinopathy with combined traction retinal detachment and rhegmatogenous retinal detachment, bilateral | E083592 | Diabetes mellitus due to underlying condition with proliferative diabetic retinopathy without macular edema, left eye | Diabetes mellitus due to underlying condition with proliferative diabetic retinopathy without macular edema, left eye | Diabetes mellitus due to underlying condition with proliferative diabetic retinopathy without macular edema, left eye | False | E113510 | | | nan | False |
| 61 | P832 | Hydrops fetalis not due to hemolytic disease | P832 | Hydrops fetalis NOS | Hydrops fetalis not due to hemolytic disease | Exact Match | True | P831 | Neonatal erythema toxicum | Neonatal erythema toxicum | Neonatal conditions | True |
| 62 | M05042 | Felty's syndrome, left hand | M05042 | Felty's syndrome, left hand | Felty's syndrome, left hand | Exact Match | True | M0502 | Felty's syndrome, elbow | | nan | True |
| 63 | S66193S | Other injury of flexor muscle, fascia and tendon of left middle finger at wrist and hand level, sequela | S56402S | Unspecified injury of extensor muscle, fascia and tendon of left index finger at forearm level, sequela | Unspecified injury of extensor muscle, fascia and tendon of left index finger at forearm level, sequela | Injury of left hand muscle, fascia and tendon, sequela | False | S66392S | Other injury of extensor muscle, fascia and tendon of right middle finger at wrist and hand level, sequela | Other injury of extensor muscle, fascia and tendon of right middle finger at wrist and hand level, sequela | Other injury of flexor muscle, fascia and tendon of left middle finger at wrist and hand level, sequela | False |
| 64 | S92023P | Displaced fracture of anterior process of unspecified calcaneus, subsequent encounter for fracture with malunion | S92025P | Nondisplaced fracture of anterior process of left calcaneus, subsequent encounter for fracture with malunion | Nondisplaced fracture of anterior process of left calcaneus, subsequent encounter for fracture with malunion | Fracture of anterior process of calcaneus, subsequent encounter for fracture with malunion | True | S92001S | Unspecified fracture of right calcaneus, sequela | Unspecified fracture of right calcaneus, sequela | Calcaneus fracture | False |
| 65 | O98519 | Other viral diseases complicating pregnancy, unspecified trimester | O9889X | | | nan | False | O98819 | Other maternal infectious and parasitic diseases complicating pregnancy, unspecified trimester | Other maternal infectious and parasitic diseases complicating pregnancy, unspecified trimester | Infections complicated pregnancy | True |

| | | | | | | | | | | | | |
|---|---|---|---|---|---|---|---|---|---|---|---|---|
| 66 | W16721S | Jumping or diving from boat striking bottom causing drowning and submersion, sequela | V9027XS | Drowning and submersion due to falling or jumping from burning water-skis, sequela | Drowning and submersion due to falling or jumping from burning water-skis, sequela | Drowning and submersion, sequela | False | W16021D | Fall into swimming pool striking bottom causing drowning and submersion, subsequent encounter | Fall into swimming pool striking bottom causing drowning and submersion, subsequent encounter | Strike pf bottom causing drowning and submersion, subsequent encounter | False |
| 67 | M86069 | Acute hematogenous osteomyelitis, unspecified tibia and fibula | M86269 | Subacute osteomyelitis, unspecified tibia and fibula | Subacute osteomyelitis, unspecified tibia and fibula | Osteomyelitis, unspecified tibia and fibula | True | M8689 | | | nan | False |
| 68 | S96891S | Other specified injury of other specified muscles and tendons at ankle and foot level, right foot, sequela | S96419S | | | nan | False | S99899S | | | nan | False |
| 69 | S82852D | Displaced trimalleolar fracture of left lower leg, subsequent encounter for closed fracture with routine healing | S82874D | Nondisplaced pilon fracture of right tibia, subsequent encounter for closed fracture with routine healing | Nondisplaced pilon fracture of right tibia, subsequent encounter for closed fracture with routine healing | Ankle fracture, subsequent encounter for closed fracture with routine healing | False | S82841K | Displaced bimalleolar fracture of right lower leg, subsequent encounter for closed fracture with nonunion | Displaced bimalleolar fracture of right lower leg, subsequent encounter for closed fracture with nonunion | Displaced right ankle fracture, subsequent encounter for closed fracture with nonunion | False |
| 70 | S8252XC | Displaced fracture of medial malleolus of left tibia, initial encounter for open fracture type IIIA, IIIB, or IIIC | S82852A | Displaced trimalleolar fracture of left lower leg, initial encounter for closed fracture | Displaced trimalleolar fracture of left lower leg, initial encounter for closed fracture | Displaced left ankle fracture, initial encounter for closed fracture | False | S82851A | Displaced trimalleolar fracture of right lower leg, initial encounter for closed fracture | Displaced trimalleolar fracture of right lower leg, initial encounter for closed fracture | Displaced ankle fracture, initial encounter for closed fracture | False |
| 71 | S4410XS | Injury of median nerve at upper arm level, unspecified arm, sequela | S14109S | Unspecified injury at unspecified level of cervical spinal cord, sequela | Unspecified injury at unspecified level of cervical spinal cord, sequela | Unspecified nerve injury of the upper body, sequela | False | S44902S | | | nan | False |
| 72 | S53136D | Medial dislocation of unspecified ulnohumeral joint, subsequent encounter | S53401D | Unspecified sprain of right elbow, subsequent encounter | Unspecified sprain of right elbow, subsequent encounter | Elbow injury | False | S53122S | Posterior subluxation of left ulnohumeral joint, sequela | Posterior subluxation of left ulnohumeral joint, sequela | Elbow injury | False |
| 73 | T8202XA | Displacement of heart valve prosthesis, initial encounter | T8201XA | Breakdown (mechanical) of heart valve prosthesis, initial encounter | Breakdown (mechanical) of heart valve prosthesis, initial encounter | Heart valve prosthetic complications | True | T827XXA | Infection and inflammatory reaction due to other cardiac and vascular devices, implants and grafts, initial encounter | Infection and inflammatory reaction due to other cardiac and vascular devices, implants and grafts, initial encounter | Heart valve prosthetic complications | False |
| 74 | S72472A | Torus fracture of lower end of left femur, initial encounter for closed fracture | S72352A | Displaced comminuted fracture of shaft of left femur, initial encounter for closed fracture | Displaced comminuted fracture of shaft of left femur, initial encounter for closed fracture | Left femur fracture, initial encounter for closed fracture | False | S72141A | Displaced intertrochanteric fracture of right femur, initial encounter for closed fracture | Displaced intertrochanteric fracture of right femur, initial encounter for closed fracture | Femur fracture, initial encounter for closed fracture | False |
| 75 | T522X4D | Toxic effect of homologues of benzene, undetermined, subsequent encounter | T5291XD | Toxic effect of unspecified organic solvent, accidental (unintentional), subsequent encounter | Toxic effect of unspecified organic solvent, accidental (unintentional), subsequent encounter | Toxic effect of substance | False | T523X5S | | | nan | False |
| 76 | D4412 | Neoplasm of uncertain behavior of left adrenal gland | D441 | Neoplasm of uncertain behavior of adrenal gland | | nan | True | D442 | Neoplasm of uncertain behavior of parathyroid gland | Neoplasm of uncertain behavior of parathyroid gland | Neoplasm of uncertain behavior | True |
| 77 | Q7101 | Congenital complete absence of right upper limb | Q7100 | Congenital complete absence of unspecified upper limb | Congenital complete absence of unspecified upper limb | Congenital absence of upper limb | True | Q7141 | Longitudinal reduction defect of right radius | Longitudinal reduction defect of right radius | Congenital right limb abnormalities | True |
| 78 | S52355D | Nondisplaced comminuted fracture of shaft of radius, left arm, subsequent encounter for closed fracture with routine healing | S52135D | Nondisplaced fracture of neck of left radius, subsequent encounter for closed fracture with routine healing | Nondisplaced fracture of neck of left radius, subsequent encounter for closed fracture with routine healing | Nondisplaced fracture of left radius, subsequent encounter for closed fracture with routine healing | False | S52311S | Greenstick fracture of shaft of radius, right arm, sequela | Greenstick fracture of shaft of radius, right arm, sequela | Radius fracture | False |
| 79 | M1A2291 | Drug-induced chronic gout, unspecified elbow, with tophus (tophi) | M1A4293 | | | nan | False | M1A00X2 | | | nan | False |
| 80 | V2501XD | Electric (assisted) bicycle driver injured in collision with railway train or railway vehicle in nontraffic accident, subsequent encounter | V190XXD | | | nan | False | V1886XS | | | nan | False |
| 81 | P093 | Abnormal findings on neonatal screening for congenital hematologic disorders | P559 | Hemolytic disease of newborn, unspecified | Hemolytic disease of newborn, unspecified | Neonatal hematologic disorders | False | P599 | Neonatal jaundice, unspecified | Neonatal jaundice, unspecified | Neonatal disorders | False |

| | | | | | | | | | | | | |
|---|---|---|---|---|---|---|---|---|---|---|---|---|
| 82 | S52241S | Displaced spiral fracture of shaft of ulna, right arm, sequela | S52215S | | | nan | False | S52312S | Greenstick fracture of shaft of radius, left arm, sequela | Greenstick fracture of shaft of radius, left arm, sequela | Lower arm fractures | False |
| 83 | S62393B | Other fracture of third metacarpal bone, left hand, initial encounter for open fracture | S62351A | Nondisplaced fracture of shaft of second metacarpal bone, left hand, initial encounter for closed fracture | Nondisplaced fracture of shaft of second metacarpal bone, left hand, initial encounter for closed fracture | Left metacarpal fracture, nitial encounter for closed fracture | False | S6234XA | | | nan | False |
| 84 | D4121 | Neoplasm of uncertain behavior of right ureter | D414 | Neoplasm of uncertain behavior of bladder | Neoplasm of uncertain behavior of bladder | Neoplasm of uncertain behavior, urinary system | True | D091 | Carcinoma in situ of other and unspecified urinary organs | | nan | False |
| 85 | I69351 | Hemiplegia and hemiparesis following cerebral infarction affecting right dominant side | I69351 | Hemiplegia and hemiparesis following cerebral infarction affecting right dominant side | Hemiplegia and hemiparesis following cerebral infarction affecting right dominant side | Exact Match | True | I69351 | Hemiplegia and hemiparesis following cerebral infarction affecting right dominant side | Hemiplegia and hemiparesis following cerebral infarction affecting right dominant side | Exact Match | True |
| 86 | S52025D | Nondisplaced fracture of olecranon process without intraarticular extension of left ulna, subsequent encounter for closed fracture with routine healing | S52612D | Displaced fracture of left ulna styloid process, subsequent encounter for closed fracture with routine healing | Displaced fracture of left ulna styloid process, subsequent encounter for closed fracture with routine healing | Left ulna fracture, subsequent encounter for closed fracture with routine healing | False | S52222S | Displaced transverse fracture of shaft of left ulna, sequela | Displaced transverse fracture of shaft of left ulna, sequela | Left ulna fracture, subsequent encounter for closed fracture with routine healing | False |
| 87 | K51519 | Left sided colitis with unspecified complications | K51812 | Other ulcerative colitis with intestinal obstruction | Other ulcerative colitis with intestinal obstruction | Colitis with complications | False | K51512 | Left sided colitis with intestinal obstruction | Left sided colitis with intestinal obstruction | Left sided colitis with complications | True |
| 88 | S52511N | Displaced fracture of right radial styloid process, subsequent encounter for open fracture type IIIA, IIIB, or IIIC with nonunion | S52133K | Displaced fracture of neck of unspecified radius, subsequent encounter for closed fracture with nonunion | Displaced fracture of neck of unspecified radius, subsequent encounter for closed fracture with nonunion | Displaced fracture of radius, subsequent encounter for fracture with nonunion | False | S52624S | | | nan | False |
| 89 | T24531D | Corrosion of first degree of right lower leg, subsequent encounter | T24621D | Corrosion of second degree of right knee, subsequent encounter | Corrosion of second degree of right knee, subsequent encounter | Corrosion of right leg, subsequent encounter | False | T24212S | Burn of second degree of left thigh, sequela | Burn of second degree of left thigh, sequela | Leg tissue injury | False |
| 90 | V162XXA | Unspecified pedal cyclist injured in collision with other nonmotor vehicle in nontraffic accident, initial encounter | V100XXA | Pedal cycle driver injured in collision with pedestrian or animal in nontraffic accident, initial encounter | Pedal cycle driver injured in collision with pedestrian or animal in nontraffic accident, initial encounter | Unspecified pedal cyclist injured in collision in nontraffic accident, initial encounter | False | V179XXA | Unspecified pedal cyclist injured in collision with fixed or stationary object in traffic accident, initial encounter | Unspecified pedal cyclist injured in collision with fixed or stationary object in traffic accident, initial encounter | Unspecified pedal cyclist injured in collision in nontraffic accident, initial encounter | False |
| 91 | C441091 | Unspecified malignant neoplasm of skin of left upper eyelid, including canthus | C44192 | Other specified malignant neoplasm of skin of right eyelid, including canthus | | nan | False | C44129 | Squamous cell carcinoma of skin of left eyelid, including canthus | | nan | True |
| 92 | T8201XD | Breakdown (mechanical) of heart valve prosthesis, subsequent encounter | T8201XD | Breakdown (mechanical) of heart valve prosthesis, subsequent encounter | Breakdown (mechanical) of heart valve prosthesis, subsequent encounter | Exact Match | True | T82795D | | | nan | False |
| 93 | V9323XA | Heat exposure on board other powered watercraft, initial encounter | X52XXXA | Prolonged stay in weightless environment, initial encounter | Prolonged stay in weightless environment, initial encounter | nan | False | 9104XA | | | nan | False |
| 94 | S0240CG | Maxillary fracture, right side, subsequent encounter for fracture with delayed healing | S0261XD | | | nan | False | S0261XD | | | nan | False |
| 95 | M7670 | Peroneal tendinitis, unspecified leg | M7660 | Achilles tendinitis, unspecified leg | Achilles tendinitis, unspecified leg | Ankle tendinitis | True | M7667 | | | nan | False |
| 96 | T2173XD | Corrosion of third degree of upper back, subsequent encounter | T2131XD | Burn of third degree of chest wall, subsequent encounter | Burn of third degree of chest wall, subsequent encounter | Third tissue injury of upper torso, subsequent encounter | False | T23328D | | | nan | False |
| 97 | V521XXS | Passenger in pick-up truck or van injured in collision with two- or three-wheeled motor vehicle in nontraffic accident, sequela | V87729S | | | nan | False | V2842XS | | | nan | False |
| 98 | S82454G | Nondisplaced comminuted fracture of shaft of right fibula, subsequent encounter for closed fracture with delayed healing | S82441D | Displaced spiral fracture of shaft of right fibula, subsequent encounter for closed fracture with routine healing | Displaced spiral fracture of shaft of right fibula, subsequent encounter for closed fracture with routine healing | Fracture of shaft of right fibula, subsequent encounter for closed fracture | False | S82445K | Nondisplaced spiral fracture of shaft of left fibula, subsequent encounter for closed fracture with nonunion | Nondisplaced spiral fracture of shaft of left fibula, subsequent encounter for closed fracture with nonunion | Fracture of shaft of fibula, subsequent encounter for closed fracture | False |

| | | | | | | | | | | | | |
|---|---|---|---|---|---|---|---|---|---|---|---|---|
| 99 | T2060XD | Corrosion of second degree of head, face, and neck, unspecified site, subsequent encounter | T2020XD | Burn of second degree of head, face, and neck, unspecified site, subsequent encounter | Burn of second degree of head, face, and neck, unspecified site, subsequent encounter | Tissue injury of second degree of head, face, and neck, unspecified site, subsequent encounter | True | T2221XD | | | nan | False |
| 100 | S52252R | Displaced comminuted fracture of shaft of ulna, left arm, subsequent encounter for open fracture type IIIA, IIIB, or IIIC with malunion | S52312K | Greenstick fracture of shaft of radius, left arm, subsequent encounter for fracture with nonunion | Greenstick fracture of shaft of radius, left arm, subsequent encounter for fracture with nonunion | Fracture of left arm, subsequent encounter | False | S52205K | | | nan | False |

Supplementary Table 3: Code-level generated outputs and similarity for ICD-10-PCS

| Case | Original ICD-10-PCS code | Original ICD-10-PCS code description | GPT-4 Code | Description matching GPT-4 code | Description matching billable GPT-4 code | GPT-4 code semantic similarity | GPT-4 code syntactic similarity | GPT-3.5 Code | Description matching GPT-3.5 code | Description matching billable GPT-3.5 code | GPT-3.5 code semantic similarity | GPT-3.5 code syntactic similarity |
|---|---|---|---|---|---|---|---|---|---|---|---|---|
| 1 | 0DW0XDZ | Revision of Intraluminal Device in Upper Intestinal Tract, External Approach | 0DJD8ZZ | Inspection of Lower Intestinal Tract, Via Natural or Artificial Opening Endoscopic | Inspection of Lower Intestinal Tract, Via Natural or Artificial Opening Endoscopic | nan | False | 0DQGXNZ | | | nan | False |
| 2 | 04HU0DZ | Insertion of Intraluminal Device into Left Peroneal Artery, Open Approach | 04LJ0DZ | Occlusion of Left External Iliac Artery with Intraluminal Device, Open Approach | Occlusion of Left External Iliac Artery with Intraluminal Device, Open Approach | Use of an intraluminal device in a lower leg artery, open approach | False | 037F04Z | Dilation of Left Hand Artery with Drug-eluting Intraluminal Device, Open Approach | Dilation of Left Hand Artery with Drug-eluting Intraluminal Device, Open Approach | Use of an intraluminal device in an artery, open approach | False |
| 3 | 041D0A7 | Bypass Left Common Iliac Artery to Left Common Iliac Artery with Autologous Arterial Tissue, Open Approach | 04C10Z3 | | | nan | False | 06H033Z | Insertion of Infusion Device into Inferior Vena Cava, Percutaneous Approach | Insertion of Infusion Device into Inferior Vena Cava, Percutaneous Approach | nan | False |
| 4 | 0TUD87Z | Supplement Urethra with Autologous Tissue Substitute, Via Natural or Artificial Opening Endoscopic | 0VQR8MZ | | | nan | False | 0UDG3ZZ | | | nan | False |
| 5 | 0SGK0KZ | Fusion of Right Tarsometatarsal Joint with Nonautologous Tissue Substitute, Open Approach | 0SGG0KZ | Fusion of Left Ankle Joint with Nonautologous Tissue Substitute, Open Approach | Fusion of Left Ankle Joint with Nonautologous Tissue Substitute, Open Approach | Fusion of foot joint with nonautologous tissue substitute, open approach | True | 0SG60JZ | Fusion of Coccygeal Joint with Synthetic Substitute, Open Approach | Fusion of Coccygeal Joint with Synthetic Substitute, Open Approach | Fusion of joint with tissue substitute, open approach | False |
| 6 | 0FU84JZ | Supplement Cystic Duct with Synthetic Substitute, Percutaneous Endoscopic Approach | 0F9R4JZ | | | nan | False | 0CJ40ZZ | | | nan | False |
| 7 | 01CK0ZZ | Extirpation of Matter from Head and Neck Sympathetic Nerve, Open Approach | 00B00ZZ | Excision of Brain, Open Approach | Excision of Brain, Open Approach | Removal of nervous system tissue in head, open approach | False | 0BH507Z | | | nan | False |
| 8 | 0SPL05Z | Removal of External Fixation Device from Left Tarsometatarsal Joint, Open Approach | 0SPJ0ZZ | | | nan | False | 0QS704Z | Reposition Left Upper Femur with Internal Fixation Device, Open Approach | Reposition Left Upper Femur with Internal Fixation Device, Open Approach | Related to use of an internal fixation device in the leg, open approach | False |
| 9 | 0YUA0JZ | Supplement Bilateral Inguinal Region with Synthetic Substitute, Open Approach | 0H9N0JZ | | | nan | False | 0JQS04Z | | | nan | False |
| 10 | 0U9C8ZZ | Drainage of Cervix, Via Natural or Artificial Opening Endoscopic | 0U9D8ZZ | | | nan | True | 0UJG8ZZ | | | nan | False |
| 11 | 0BC63ZZ | Extirpation of Matter from Right Lower Lobe Bronchus, Percutaneous Approach | 0B9M3ZZ | Drainage of Bilateral Lungs, Percutaneous Approach | Drainage of Bilateral Lungs, Percutaneous Approach | Removal of material from lungs, percutaneous approach | False | 0WUJXZZ | | | nan | False |
| 12 | CP2G1ZZ | Tomographic (Tomo) Nuclear Medicine Imaging of Thoracic Spine using Technetium 99m (Tc-99m) | BW24ZZZ | Computerized Tomography (CT Scan) of Chest and Abdomen | Computerized Tomography (CT Scan) of Chest and Abdomen | Chest imaging | False | 04NJ3ZZ | Release Left External Iliac Artery, Percutaneous Approach | Release Left External Iliac Artery, Percutaneous Approach | nan | False |
| 13 | 0N8J3ZZ | Division of Left Lacrimal Bone, Percutaneous Approach | 02PU3ZZ | | | nan | False | 0SG107Z | | | nan | False |
| 14 | 0F9630Z | Drainage of Left Hepatic Duct with Drainage Device, Percutaneous Approach | 0F993DZ | | | nan | False | 0D5G7DZ | | | nan | False |
| 15 | 09B14ZX | Excision of Left External Ear, Percutaneous Endoscopic Approach, Diagnostic | 0CBP4ZX | | | nan | False | 0HBV4ZZ | | | nan | False |
| 16 | 0RJTXZZ | Inspection of Left Carpometacarpal Joint, External Approach | 0SQ50ZZ | Repair Sacrococcygeal Joint, Open Approach | Repair Sacrococcygeal Joint, Open Approach | nan | False | 0JVS8ZZ | | | nan | False |
| 17 | 047Q0Z1 | Dilation of Left Anterior Tibial Artery using Drug-Coated Balloon, Open Approach | 04V30DZ | Restriction of Hepatic Artery with Intraluminal Device, Open Approach | Restriction of Hepatic Artery with Intraluminal Device, Open Approach | nan | False | 02762ZZ | | | nan | False |

| | | | | | | | | | | | | |
|---|---|---|---|---|---|---|---|---|---|---|---|---|
| 18 | 0HRX3JZ | Replacement of Left Nipple with Synthetic Substitute, Percutaneous Approach | 0HRPXJZ | | | nan | False | 0WJF3ZZ | Inspection of Abdominal Wall, Percutaneous Approach | Inspection of Abdominal Wall, Percutaneous Approach | nan | False |
| 19 | 0D1B0JB | Bypass Ileum to Ileum with Synthetic Substitute, Open Approach | 0DBP0JZ | | | nan | False | 06UK3ZZ | | | nan | False |
| 20 | 019L3ZZ | Drainage of Thoracic Sympathetic Nerve, Percutaneous Approach | 00P03ZZ | | | nan | False | 0BCK4ZZ | Extirpation of Matter from Right Lung, Percutaneous Endoscopic Approach | Extirpation of Matter from Right Lung, Percutaneous Endoscopic Approach | Removal of material from thorax, percutaneous approach | False |
| 21 | B31P1ZZ | Fluoroscopy of Thoraco-Abdominal Aorta using Low Osmolar Contrast | BW28ZZZ | Computerized Tomography (CT Scan) of Head | Computerized Tomography (CT Scan) of Head | Both are imaging techniques | False | 0RFV0JZ | | | nan | False |
| 22 | 04NF3ZZ | Release Left Internal Iliac Artery, Percutaneous Approach | 04U03ZZ | | | nan | False | 0RBC0ZZ | Excision of Right Temporomandibular Joint, Open Approach | Excision of Right Temporomandibular Joint, Open Approach | nan | False |
| 23 | 07924ZZ | Drainage of Left Neck Lymphatic, Percutaneous Endoscopic Approach | 0W9P4ZZ | | | nan | False | 0DPG3ZZ | | | nan | False |
| 24 | 0RG83KJ | Fusion of 8 or more Thoracic Vertebral Joints with Nonautologous Tissue Substitute, Posterior Approach, Anterior Column, Percutaneous Approach | 0RGX0KZ | Fusion of Left Finger Phalangeal Joint with Nonautologous Tissue Substitute, Open Approach | Fusion of Left Finger Phalangeal Joint with Nonautologous Tissue Substitute, Open Approach | Fusion of joint with nonautologous tissue substitute | False | 0Q9C07Z | | | nan | False |
| 25 | 3E09XKZ | Introduction of Other Diagnostic Substance into Nose, External Approach | 3E0LX3Z | | | nan | False | 0WQNXZZ | Repair Female Perineum, External Approach | Repair Female Perineum, External Approach | nan | False |
| 26 | 0YJ94ZZ | Inspection of Right Lower Extremity, Percutaneous Endoscopic Approach | 0W9H4ZZ | Drainage of Retroperitoneum, Percutaneous Endoscopic Approach | Drainage of Retroperitoneum, Percutaneous Endoscopic Approach | nan | False | 0JVG8ZZ | | | nan | False |
| 27 | 0UQ14ZZ | Repair Left Ovary, Percutaneous Endoscopic Approach | 0UT94ZZ | Resection of Uterus, Percutaneous Endoscopic Approach | Resection of Uterus, Percutaneous Endoscopic Approach | Female reproductive organ surgery, percutaneous endoscopic approach | False | 0U5H38Z | | | nan | False |
| 28 | DD17B8Z | Low Dose Rate (LDR) Brachytherapy of Rectum using Iridium 192 (Ir-192) | D1HW0ZZ | | | nan | False | 0KQG0Z2 | | | nan | False |
| 29 | 047L4Z1 | Dilation of Left Femoral Artery using Drug-Coated Balloon, Percutaneous Endoscopic Approach | 04V34DZ | Restriction of Hepatic Artery with Intraluminal Device, Percutaneous Endoscopic Approach | Restriction of Hepatic Artery with Intraluminal Device, Percutaneous Endoscopic Approach | nan | False | 027N3ZZ | | | Left Femoral Artery | False |
| 30 | 02710GZ | Dilation of Coronary Artery, Two Arteries with Four or More Intraluminal Devices, Open Approach | 02703DZ | Dilation of Coronary Artery, One Artery with Intraluminal Device, Percutaneous Approach | Dilation of Coronary Artery, One Artery with Intraluminal Device, Percutaneous Approach | Dilation of coronary artery with intraluminal device | False | 021B04Z | | | nan | False |
| 31 | 00XM4ZS | Transfer Facial Nerve to Hypoglossal Nerve, Percutaneous Endoscopic Approach | 00CN4ZZ | Extirpation of Matter from Acoustic Nerve, Percutaneous Endoscopic Approach | Extirpation of Matter from Acoustic Nerve, Percutaneous Endoscopic Approach | Cranial nerve surgery, percutaneous endoscopic approach | False | 0J1D8NZ | | | nan | False |
| 32 | 0QUR0JZ | Supplement Left Toe Phalanx with Synthetic Substitute, Open Approach | 0SP60JZ | Removal of Synthetic Substitute from Coccygeal Joint, Open Approach | Removal of Synthetic Substitute from Coccygeal Joint, Open Approach | Related to synthetic substitute, open approach | False | 0SP504Z | Removal of Internal Fixation Device from Sacrococcygeal Joint, Open Approach | Removal of Internal Fixation Device from Sacrococcygeal Joint, Open Approach | nan | False |
| 33 | 04VC3EZ | Restriction of Right Common Iliac Artery with Branched or Fenestrated Intraluminal Device, One or Two Arteries, Percutaneous Approach | 047K3DZ | Dilation of Right Femoral Artery with Intraluminal Device, Percutaneous Approach | Dilation of Right Femoral Artery with Intraluminal Device, Percutaneous Approach | Use of an intraluminal device in a upper leg artery, percutaneous approach | False | 0RB408Z | | | nan | False |
| 34 | 04UJ4JZ | Supplement Left External Iliac Artery with Synthetic Substitute, Percutaneous Endoscopic Approach | 04WQ4JZ | | | nan | False | 0VJ707Z | | | nan | False |
| 35 | D91D99Z | High Dose Rate (HDR) Brachytherapy of Nasopharynx using Iodine 125 (I-125) | D1H30ZZ | | | nan | False | 0Q4D0ZZ | | | nan | False |

| | | | | | | | | | | | | |
|---|---|---|---|---|---|---|---|---|---|---|---|---|
| 36 | 0PS93ZZ | Reposition Right Clavicle, Percutaneous Approach | 0PS03ZZ | Reposition Sternum, Percutaneous Approach | Reposition Sternum, Percutaneous Approach | Reposition upper torso bone, percutaneous approach | True | 0R3R0NZ | | | nan | False |
| 37 | 00XK0ZN | Transfer Trigeminal Nerve to Acoustic Nerve, Open Approach | 00C10ZZ | Extirpation of Matter from Cerebral Meninges, Open Approach | Extirpation of Matter from Cerebral Meninges, Open Approach | Central nervous system surgery, open approach | False | 0XQD0ZZ | Repair Right Lower Arm, Open Approach | Repair Right Lower Arm, Open Approach | nan | False |
| 38 | HZ87ZZZ | Medication Management for Substance Abuse Treatment, Bupropion | HZ2C4ZZ | | | nan | False | 3E0D30Z | | | nan | False |
| 39 | 0MBG4ZX | Excision of Rib(s) Bursa and Ligament, Percutaneous Endoscopic Approach, Diagnostic | 0WBQ4ZX | | | nan | False | 0R1R04Z | | | nan | False |
| 40 | 02H00YZ | Insertion of Other Device into Coronary Artery, One Artery, Open Approach | 02700ZZ | Dilation of Coronary Artery, One Artery, Open Approach | Dilation of Coronary Artery, One Artery, Open Approach | Coronary artery | False | 021R0ZZ | | | Coronary Artery | False |
| 41 | 0BW0X2Z | Revision of Monitoring Device in Tracheobronchial Tree, External Approach | 0BJD8ZZ | | | nan | False | 0BH98ZZ | | | nan | False |
| 42 | 039C0ZZ | Drainage of Left Radial Artery, Open Approach | 03CJ0ZZ | Extirpation of Matter from Left Common Carotid Artery, Open Approach | Extirpation of Matter from Left Common Carotid Artery, Open Approach | Removal of material from left-sided arteries, open approach | False | 0J8H38Z | | | nan | False |
| 43 | F0701UZ | Muscle Performance Treatment of Neurological System - Head and Neck using Prosthesis | HZ8C5ZZ | | | nan | False | 5A135LN | | | nan | False |
| 44 | 30243W0 | Transfusion of Autologous Factor IX into Central Vein, Percutaneous Approach | 30233Y0 | Transfusion of Autologous Hematopoietic Stem Cells into Peripheral Vein, Percutaneous Approach | Transfusion of Autologous Hematopoietic Stem Cells into Peripheral Vein, Percutaneous Approach | Transfusion of autologous substances into venous system, percutaneous approach | False | 3E043WZ | | | nan | False |
| 45 | 0VW407Z | Revision of Autologous Tissue Substitute in Prostate and Seminal Vesicles, Open Approach | 0VQW0ZZ | | | nan | False | 0S2H8Z2 | | | nan | False |
| 46 | 057B3DZ | Dilation of Right Basilic Vein with Intraluminal Device, Percutaneous Approach | 05V23DZ | | | nan | False | 037U0ZZ | Dilation of Right Thyroid Artery, Open Approach | Dilation of Right Thyroid Artery, Open Approach | Dilation of vessel | False |
| 47 | 0NT70ZZ | Resection of Occipital Bone, Open Approach | 01BN0ZZ | Excision of Lumbar Sympathetic Nerve, Open Approach | Excision of Lumbar Sympathetic Nerve, Open Approach | nan | False | 0HBU07Z | | | nan | False |
| 48 | 0LRD4KZ | Replacement of Left Thorax Tendon with Nonautologous Tissue Substitute, Percutaneous Endoscopic Approach | 0KQQ4KZ | | | nan | False | 0QH104Z | Insertion of Internal Fixation Device into Sacrum, Open Approach | Insertion of Internal Fixation Device into Sacrum, Open Approach | nan | False |
| 49 | 0VUG8KZ | Supplement Left Spermatic Cord with Nonautologous Tissue Substitute, Via Natural or Artificial Opening Endoscopic | 0VBQ8KZ | | | nan | False | 0VJF7ZZ | | | nan | False |
| 50 | 0TWB4YZ | Revision of Other Device in Bladder, Percutaneous Endoscopic Approach | 0TTD4ZZ | Resection of Urethra, Percutaneous Endoscopic Approach | Resection of Urethra, Percutaneous Endoscopic Approach | Urinary tract surgery, percutaneous endoscopic approach | False | 0PBG04Z | | | nan | False |
| 51 | CW13YZZ | Planar Nuclear Medicine Imaging of Chest using Other Radionuclide | C0210ZZ | | | nan | False | B32Z1ZZ | | | nan | False |
| 52 | 037A4EZ | Dilation of Left Ulnar Artery with Two Intraluminal Devices, Percutaneous Endoscopic Approach | 027034Z | Dilation of Coronary Artery, One Artery with Drug-eluting Intraluminal Device, Percutaneous Approach | Dilation of Coronary Artery, One Artery with Drug-eluting Intraluminal Device, Percutaneous Approach | Dilation of artery with intraluminal device | False | 02703JZ | | | nan | False |
| 53 | 0TNC7ZZ | Release Bladder Neck, Via Natural or Artificial Opening | 0T9V7ZZ | | | nan | False | 0V5L0ZZ | Destruction of Bilateral Epididymis, Open Approach | Destruction of Bilateral Epididymis, Open Approach | Genitourinary surgery | False |
| 54 | 0THC0LZ | Insertion of Artificial Sphincter into Bladder Neck, Open Approach | 0TH10HZ | | | nan | False | 0W8J0ZZ | | | nan | False |

| | | | | | | | | | | | | |
|---|---|---|---|---|---|---|---|---|---|---|---|---|
| 55 | 0V934ZZ | Drainage of Bilateral Seminal Vesicles, Percutaneous Endoscopic Approach | 0T9B8ZZ | Drainage of Bladder, Via Natural or Artificial Opening Endoscopic | Drainage of Bladder, Via Natural or Artificial Opening Endoscopic | Drainage of genitourinary structures | False | 0THT7ZZ | | | nan | False |
| 56 | 0JHD0NZ | Insertion of Tissue Expander into Right Upper Arm Subcutaneous Tissue and Fascia, Open Approach | 0HRBXJZ | Replacement of Right Upper Arm Skin with Synthetic Substitute, External Approach | Replacement of Right Upper Arm Skin with Synthetic Substitute, External Approach | Manipulation of right upper arm tissue | False | 0DQ67ZZ | Repair Stomach, Via Natural or Artificial Opening | Repair Stomach, Via Natural or Artificial Opening | nan | False |
| 57 | 0DC58ZZ | Extirpation of Matter from Esophagus, Via Natural or Artificial Opening Endoscopic | 0DB88ZZ | Excision of Small Intestine, Via Natural or Artificial Opening Endoscopic | Excision of Small Intestine, Via Natural or Artificial Opening Endoscopic | Upper gastrointestinal endoscopic procedures | False | 0DJ98ZZ | | | nan | False |
| 58 | 0W9240Z | Drainage of Face with Drainage Device, Percutaneous Endoscopic Approach | 0H9A8KZ | | | nan | False | 0S9J7ZZ | | | nan | False |
| 59 | 051947Y | Bypass Right Brachial Vein to Upper Vein with Autologous Tissue Substitute, Percutaneous Endoscopic Approach | 051V3DZ | | | nan | False | 06T80ZZ | | | nan | False |
| 60 | 0YQTXZZ | Repair Right 3rd Toe, External Approach | 0SPC0ZZ | | | nan | False | 0YQS0ZZ | Repair Left 2nd Toe, Open Approach | Repair Left 2nd Toe, Open Approach | Toe repair, open approach | False |
| 61 | B320Z2Z | Computerized Tomography (CT Scan) of Thoracic Aorta using Intravascular Optical Coherence | BP32ZZZ | | | nan | False | B231ZZZ | Magnetic Resonance Imaging (MRI) of Multiple Coronary Arteries | Magnetic Resonance Imaging (MRI) of Multiple Coronary Arteries | Imaging of chest arteries | False |
| 62 | 0QHB38Z | Insertion of Limb Lengthening External Fixation Device into Right Lower Femur, Percutaneous Approach | 0QS604Z | Reposition Right Upper Femur with Internal Fixation Device, Open Approach | Reposition Right Upper Femur with Internal Fixation Device, Open Approach | Right femur surgery involving fixation device | False | 0QWB8ZZ | | | nan | False |
| 63 | 07544ZZ | Destruction of Left Upper Extremity Lymphatic, Percutaneous Endoscopic Approach | 07L83ZZ | Occlusion of Right Internal Mammary Lymphatic, Percutaneous Approach | Occlusion of Right Internal Mammary Lymphatic, Percutaneous Approach | Obstruction of upper lymphatic structures | False | 04T12JZ | | | nan | False |
| 64 | 04VT4ZZ | Restriction of Right Peroneal Artery, Percutaneous Endoscopic Approach | 04L23DZ | Occlusion of Gastric Artery with Intraluminal Device, Percutaneous Approach | Occlusion of Gastric Artery with Intraluminal Device, Percutaneous Approach | Blocking arteries | False | 02720NZ | | | nan | False |
| 65 | 0RGP07Z | Fusion of Left Wrist Joint with Autologous Tissue Substitute, Open Approach | 0RG10KZ | | | nan | False | 0RQF0JZ | | | nan | False |
| 66 | 08NE3ZZ | Release Right Retina, Percutaneous Approach | 08P04ZZ | | | nan | False | 0LQ50ZZ | Repair Right Lower Arm and Wrist Tendon, Open Approach | Repair Right Lower Arm and Wrist Tendon, Open Approach | nan | False |
| 67 | 0KXK0Z1 | Transfer Right Abdomen Muscle with Subcutaneous Tissue, Open Approach | 0HRG0ZZ | | | nan | False | 0KQG0ZZ | Repair Left Trunk Muscle, Open Approach | Repair Left Trunk Muscle, Open Approach | Manipulation of abdominal muscles, open approach | False |
| 68 | 0QNJ0ZZ | Release Right Fibula, Open Approach | 0PQ70ZZ | Repair Right Glenoid Cavity, Open Approach | Repair Right Glenoid Cavity, Open Approach | nan | False | 0QS605Z | Reposition Right Upper Femur with External Fixation Device, Open Approach | Reposition Right Upper Femur with External Fixation Device, Open Approach | Right leg skeletal repair | False |
| 69 | 04WYX3Z | Revision of Infusion Device in Lower Artery, External Approach | 02W30ZZ | | | nan | False | 031D4ZZ | | | nan | False |
| 70 | 0BV60CZ | Restriction of Right Lower Lobe Bronchus with Extraluminal Device, Open Approach | 0B513DZ | | | nan | False | 0QBB3ZZ | Excision of Right Lower Femur, Percutaneous Approach | Excision of Right Lower Femur, Percutaneous Approach | nan | False |
| 71 | 0WP90JZ | Removal of Synthetic Substitute from Right Pleural Cavity, Open Approach | 0B9H0KZ | | | nan | False | 0TJ50ZZ | Inspection of Kidney, Open Approach | Inspection of Kidney, Open Approach | nan | False |
| 72 | 0JB50ZZ | Excision of Left Neck Subcutaneous Tissue and Fascia, Open Approach | 0HB60ZZ | | | nan | False | 0JQ70ZZ | Repair Back Subcutaneous Tissue and Fascia, Open Approach | Repair Back Subcutaneous Tissue and Fascia, Open Approach | Involving subcutaneous Tissue and Fascia, Open Approach | False |

| | | | | | | | | | | | | |
|---|---|---|---|---|---|---|---|---|---|---|---|---|
| 73 | 0D1M4JP | Bypass Descending Colon to Rectum with Synthetic Substitute, Percutaneous Endoscopic Approach | 0D1C3DZ | | | nan | False | 06P84ZZ | | | nan | False |
| 74 | 0D1B8ZP | Bypass Ileum to Rectum, Via Natural or Artificial Opening Endoscopic | 0D1B8DZ | | | nan | False | 0DVG4ZZ | Restriction of Left Large Intestine, Percutaneous Endoscopic Approach | Restriction of Left Large Intestine, Percutaneous Endoscopic Approach | nan | False |
| 75 | HZ48ZZZ | Group Counseling for Substance Abuse Treatment, Confrontational | HZ2ZZZZ | Detoxification Services for Substance Abuse Treatment | Detoxification Services for Substance Abuse Treatment | nan | False | 9A645GZ | | | nan | False |
| 76 | 0BV94ZZ | Restriction of Lingula Bronchus, Percutaneous Endoscopic Approach | 0B513ZZ | Destruction of Trachea, Percutaneous Approach | Destruction of Trachea, Percutaneous Approach | Obstructing portions of the respiratory tract | False | 0BQ37NZ | | | nan | False |
| 77 | 0D120ZB | Bypass Middle Esophagus to Ileum, Open Approach | 0D1M0DZ | | | nan | False | 0DTG4ZZ | Resection of Left Large Intestine, Percutaneous Endoscopic Approach | Resection of Left Large Intestine, Percutaneous Endoscopic Approach | nan | False |
| 78 | 0D7A8DZ | Dilation of Jejunum with Intraluminal Device, Via Natural or Artificial Opening Endoscopic | 0D548ZZ | Destruction of Esophagogastric Junction, Via Natural or Artificial Opening Endoscopic | Destruction of Esophagogastric Junction, Via Natural or Artificial Opening Endoscopic | Endoscopic procedures in the gastrointestinal tract | False | 0WQH4ZZ | | | nan | False |
| 79 | 0HWT7KZ | Revision of Nonautologous Tissue Substitute in Right Breast, Via Natural or Artificial Opening | 0HQ78KZ | | | nan | False | 0HBT0ZX | Excision of Right Breast, Open Approach, Diagnostic | Excision of Right Breast, Open Approach, Diagnostic | Involving right breast | False |
| 80 | 07PK43Z | Removal of Infusion Device from Thoracic Duct, Percutaneous Endoscopic Approach | 06S73DZ | | | nan | False | 06T20ZZ | | | nan | False |
| 81 | 03703ZZ | Dilation of Right Internal Mammary Artery, Percutaneous Approach | 02720ZZ | Dilation of Coronary Artery, Three Arteries, Open Approach | Dilation of Coronary Artery, Three Arteries, Open Approach | Dilation of chest artery | False | 02RG0ZZ | | | nan | False |
| 82 | 03VM4HZ | Restriction of Right External Carotid Artery with Intraluminal Device, Flow Diverter, Percutaneous Endoscopic Approach | 031C3DZ | | | nan | False | 02721CZ | | | nan | False |
| 83 | 03RD4KZ | Replacement of Right Hand Artery with Nonautologous Tissue Substitute, Percutaneous Endoscopic Approach | 04S63DZ | | | nan | False | 027H0NZ | | | nan | False |
| 84 | 06R20JZ | Replacement of Gastric Vein with Synthetic Substitute, Open Approach | 06270KZ | | | nan | False | 0V597ZZ | | | nan | False |
| 85 | DD14BB1 | Low Dose Rate (LDR) Brachytherapy of Ileum using Palladium 103 (Pd-103), Unidirectional Source | D3314UZ | | | nan | False | 0TC23ZZ | | | nan | False |
| 86 | 03B53ZX | Excision of Right Axillary Artery, Percutaneous Approach, Diagnostic | 03504ZZ | Destruction of Right Internal Mammary Artery, Percutaneous Endoscopic Approach | Destruction of Right Internal Mammary Artery, Percutaneous Endoscopic Approach | Damaging right upper body arteries | False | 0RPY04Z | | | nan | False |
| 87 | 0MMS0ZZ | Reattachment of Right Foot Bursa and Ligament, Open Approach | 0SP90ZZ | | | nan | False | 0QY20ZZ | | | nan | False |
| 88 | 03U33JZ | Supplement Right Subclavian Artery with Synthetic Substitute, Percutaneous Approach | 04120KZ | | | nan | False | 02770JZ | | | nan | False |
| 89 | 03CN3Z7 | Extirpation of Matter from Left External Carotid Artery using Stent Retriever, Percutaneous Approach | 03104DZ | | | nan | False | 02743NZ | | | nan | False |
| 90 | 06184Z9 | Bypass Portal Vein to Right Renal Vein, Percutaneous Endoscopic Approach | 02S43ZZ | | | nan | False | 06N93ZZ | Release Right Renal Vein, Percutaneous Approach | Release Right Renal Vein, Percutaneous Approach | Involving right renal vein | False |

| | | | | | | | | | | | | |
|---|---|---|---|---|---|---|---|---|---|---|---|---|
| 91 | 03UV47Z | Supplement Left Thyroid Artery with Autologous Tissue Substitute, Percutaneous Endoscopic Approach | 041C3KZ | | | nan | False | 02790NZ | | | nan | False |
| 92 | 0T974ZZ | Drainage of Left Ureter, Percutaneous Endoscopic Approach | 0T998ZZ | | | nan | False | 0TGU7ZZ | | | nan | False |
| 93 | 031409W | Bypass Left Subclavian Artery to Lower Extremity Vein with Autologous Venous Tissue, Open Approach | 041C0JZ | | | nan | False | 0QSF0JZ | | | nan | False |
| 94 | 0F190Z4 | Bypass Common Bile Duct to Stomach, Open Approach | 0F194DZ | | | nan | False | 0DJD0ZZ | Inspection of Lower Intestinal Tract, Open Approach | Inspection of Lower Intestinal Tract, Open Approach | Related to the gastrointestinal tract | False |
| 95 | 009900Z | Drainage of Thalamus with Drainage Device, Open Approach | 01HY0KZ | | | nan | False | 0TGK7ZZ | | | nan | False |
| 96 | 089Q3ZZ | Drainage of Right Lower Eyelid, Percutaneous Approach | 08HR4ZZ | | | nan | False | 0LQM0ZZ | Repair Left Upper Leg Tendon, Open Approach | Repair Left Upper Leg Tendon, Open Approach | nan | False |
| 97 | 0S9430Z | Drainage of Lumbosacral Disc with Drainage Device, Percutaneous Approach | 05HY3KZ | | | nan | False | 0RG74ZZ | | | nan | False |
| 98 | 05QN3ZZ | Repair Left Internal Jugular Vein, Percutaneous Approach | 03RC3ZZ | | | nan | False | 0RNB4ZZ | Release Thoracolumbar Vertebral Disc, Percutaneous Endoscopic Approach | Release Thoracolumbar Vertebral Disc, Percutaneous Endoscopic Approach | nan | False |
| 99 | 0MBN0ZZ | Excision of Right Knee Bursa and Ligament, Open Approach | 0K9H0ZZ | Drainage of Right Thorax Muscle, Open Approach | Drainage of Right Thorax Muscle, Open Approach | nan | False | 0QY90ZZ | | | nan | False |
| 100 | 047R46Z | Dilation of Right Posterior Tibial Artery with Three Drug-eluting Intraluminal Devices, Percutaneous Endoscopic Approach | 027F34Z | Dilation of Aortic Valve with Drug-eluting Intraluminal Device, Percutaneous Approach | Dilation of Aortic Valve with Drug-eluting Intraluminal Device, Percutaneous Approach | Dilation of vascular structure with drug-eluting intraluminal devices | False | 027S0JZ | | | nan | False |
